# The Coronavirus Disease (COVID-19) Pandemic: Simulation-Based Assessment of Outbreak Responses and Post Peak Strategies

**DOI:** 10.1101/2020.04.13.20063610

**Authors:** Jeroen Struben

## Abstract

It is critical to understand the impact of distinct policy interventions to the ongoing coronavirus disease pandemic. I develop a flexible behavioral, dynamic, and sectorial epidemic policy model comprising both endogenous virus transmission and public health and citizen responses. Applicable to the full epidemic cycle including confinement, deconfinement, and resurgence, the model allows exploring the multivariate impact of distinct policy interventions, including general and targeted testing and social contact reduction efforts. Using a cross-sectional calibration to data on the ongoing coronavirus disease outbreak about reported cases and deaths, tests performed, and social interactions from six countries (South Korea, Germany, Italy, France, Sweden, and the United States), I demonstrate how early, rapid, and extensive buildup of testing and social contact reduction efforts interplay to suppress the outbreak. I then use the model to show and quantify limits to the extent of deconfinement and illustrate the critical role of targeted approaches for managing post peak deconfinement and resurgence. To aid necessary public and expert understanding of outbreak control strategies the model is accessible in the form of a web-based management flight simulator.

## Introduction

On March 11 the World Health Organization declared the coronavirus disease (COVID-19) outbreak a global pandemic (WHO, 2020a). As of mid-May over 4 Million cases of COVID-19 virus infections and over 300 Thousand deaths have been reported (Roser, Ritchie, and Ortiz-Ospina, 2020). While the disease has affected over 200 countries, areas, and territories, outbreak patterns and responses have varied widely (Cohen and Kupferschmidt, 2020). In Singapore, extensive restrictions on movement started three days after the first discovered case (Xianbai, 2020), whereas other countries have been slower to reduce social and economic interactions. South Korea deployed rapid and large-scale testing across the population, while the United States has initially been slow to build up testing capacity (Cho, 2020). Across Europe, responses have also differed considerably (Politico, 2020). On March 16 the French government ordered citizens to stay at home except for essential activities (Erlanger, 2020). In contrast, the UK government initially suggested avoidance of public places (Triggle 2020), yet bars, restaurants, and museums remained open. Prime Minister Boris Johnson subsequently changed course, but confusion persisted over what was and wasn’t allowed (Mason 2020). The same virus and disease, different government and citizen responses.

Early data on the developing pandemic (Roser et al. 2020) suggested the vital importance of rapid scaling up of testing and interventions aimed at reducing physically proximate interactions (social contacts) for decreasing total and peak infections. But, these different approaches and pathways across countries raise a number of critical questions, including: How do these various interventions impact the outbreak patterns? Why is large-scale and early testing so important? What is the differential effect between enforcing or discouraging social contact reduction? How sensitive are interventions to imperfect compliance by citizens? How do different interventions interact to alter outbreak patterns? How much can interventions relax once the outbreak is under control? How important are targeted interventions? And, what is the importance of coordinating and aligning efforts across countries? Answering such questions requires detailed understanding of the diffusion patterns of infectious diseases.

The central purpose of the behavioral infectious disease policy model I develop here is to analyze the individual and joint impact of diverse specific public health control measures over the epidemic cycle including first outbreak response, first-wave deconfinement, and virus resurgence for addressing such questions. Achieving this requires capturing critical virus and transmission characteristics, including transmission rates, and incubation and infectious periods. For this reason, the model follows the principles of Susceptible-Infected-Recovered (SIR) compartment models (Brauer, Castillo-Chavez, and Castillo-Chavez, 2012), a widely used class of epidemiological models specifically designed for studying these virus transmission dynamics. Second, the model must represent the role of human responses to the outbreak and how these in turns alter the outbreak dynamics (Ferguson et al. 2020a). To do this, the model follows principles of behavioral dynamic modeling (Sterman 2001) in line with a long history of compartment-based infectious disease models within the field of system dynamics (Thompson and Tebbens. 2008; Darabi and Hosseinichimeh, 2020) with models developed for studying issues ranging from pharmaceutical effectiveness in HIV/AIDS (Dangerfield et al., 2001), psychological factors in social contact adjustment in Ebola (Pruyt et al. 2015), to endgame strategies in the context of the poliovirus (Thompson and Tebbens 2007; Duintjer Tebbens et al., 2005).

Yet, while building on these two streams of research, policy relevance demands particular attention to key issues central to the specific outbreak. The present COVID-19 outbreak is characterized by high first-wave virus transmission rates, by largely undetected spreading - in part due to the dominance of low-symptom cases - but relatively high death rates (Ferguson et al. 2020; Kissler et al. 2020a). Further, presently, there is much uncertainty about timelines of vaccination development and deployment and duration of immunity (Kissler et al. 2020a). Therefore, for example, beyond endgame strategies, managing the COVID-19 outbreak requires effective nonpharmaceutical strategies including for deconfinement and outbreak resurgence. Interventions that effectively do so need to be grounded in deep understanding of the dynamic complexities resulting from the different ways by which, in response to the present outbreak, populations may alter their social contacts, health experts expand testing and reporting, and policy makers implement social distancing and surveillance policies, and from their collective impact on the outbreak.

To capture first-wave and resurgence dynamics for COVID-19 the model includes endogenous social distancing responses by citizens and interventions by policy makers as well as the buildup and execution of testing and reporting. Specifically, the core of the model forms a “susceptible exposed infectious recovered” (SEIR) model (Hetchote 2000). Importantly, within the model, as in real life, citizens and policy makers respond to reported data of tested– not actual – positive cases during a progressing virus outbreak. Testing capacity, initially non-existent, has to be built-up. Further, the model differentiates mild cases –exhibiting mild symptoms or being fully asymptomatic – from severe cases. Because the former group is generally less likely to be tested but nevertheless infectious, this differentiation affects case detection and citizen responses, and therefore the overall outbreak patterns. The model explicitly captures both reactive (based on the exhibition of symptoms) and proactive testing (based on field testing) approaches as well as different types of interventions including general population social distancing, home confinement of suspected cases, and quarantining of positive (detected) cases. The model structure has some generality in the sense that it can be used vary assumptions about virus transmission parameters, citizen and policy behavior, and demographic characteristics, within and across socio-demographic segments.^1^ The model tracks key epidemic variables over time (including the population within the various epidemic stages and the reproductive number - the average number of secondary cases that one case generates over the course of its infectious period), as well as clinical (hospitalizations, deaths) and behavioral variables (degree of social contacts, and home-confined population, reported versus actual cases, etc.).

Here I use the model to examine effectiveness of interventions to manage different phases of the COVID-19 pandemic. To do this, in a first experiment, I develop a Baseline simulation run that calibrates the model against the rapidly developing data and literature on the ongoing outbreak (Roser, Ritchie and Ortiz-Ospina, 2020; Dong, Du, and Gardner, 2020; The Lancet, 2020). The Baseline involves 6 different countries with varying outbreak paths and intervention choices (South Korea, Germany, Italy, France, Sweden, and the United States) with parameters estimated through a cross-sectional calibration to data on reported cases and deaths, tests performed, and social contacts. I then use the calibrated model to perform analyses within four additional experiments. First, I examine sensitivity of outcomes to individual and multiple outcomes to policy responses. Next I examine, within a more stylized context, efforts and timing to deconfine and interventions that help reduce virus resurgence. In the final experiment, motivated by the large variation in clinical outcomes across countries, I examine in more depth the impact of heterogeneity in socio-demographic (eg age) related vulnerability to the virus on overall outbreak.

The analysis results shows, first, how timing and extensiveness of testing and of social contact reduction measures interplay and can explain a great share of the differences in outbreak pathways across countries. More quantitatively, while heeding limitations in confidence in some of the specific parameter values and trajectories at this point during the outbreak, sensitivity analysis in the case of the United States suggests that a one-week earlier (later) ramping up of either testing capacity buildup or of social distancing action could have led to on the order of 50% fewer (additional) actual deaths. These large effects result in part because of interactive effects between strong coupled positive feedback loops. For example, testing efforts do not only allow case identification and isolation but also help, due to increased awareness of the outbreak, accelerate interventions aimed at general contact reductions. Second, I show how, once countries fall behind in curtailing the outbreak, it is difficult to catch up. Once the gap between tested and actual cases expands, a positive feedback involving transmissions from unidentified cases drives detection interventions towards late-stage reactive – rather than early-stage proactive - case detection. Third, in terms of deconfinement strategies, I show that, as long as no pharmaceutical solutions are available at scale, risks of renewed large-scale outbreaks are very high when social contact rates exceed 60-70% of the pre-pandemic levels. Those risk depend importantly on a country’s preparedness for deconfinement as well as on confinement policies prior to deconfinement. Specifically, the presence of effective and extensive targeted testing and intervention approaches (contact tracing and testing, broader suspect case isolation, surveillance, and their compliance) are vital to reduce the multifaceted risks and impacts of resurgence. Fourth, as a more general finding, the model allows pinpointing the origins of the fundamentally complex dynamics of infectious diseases caused by both virus transmission and human behavior. Powerful positive feedbacks that accelerate infections combine with delays in infection detectability, inertia in the buildup of testing capabilities, and with challenges in rapidly limiting human contacts. In capturing responses behaviorally and including important delays and interdependencies the model further helps understand why and how swift and comprehensive responses can reduce the impact of epidemic outbreaks. In clarifying these endogenous dynamics, our model provides insights that are fundamentally different from and complement to policy models that study interventions as exogenous shocks (Kissler et al. 2020b). By allowing the exploration of different intervention strategies through endogenous behavior - from social distancing advice, to home confinement of suspect cases, we provide a general quantitative framework for better understand under what conditions what combinations of measures help successful reversing epidemic outbreak – either at an early stage of an outbreak or during the later stage of resurgence management.

Finally, because much of the success depends on collective involvement from not only experts, but also from policy makers shaping policies, volunteers with efforts within communities, citizen compliance, and media communication that all need to better understand these dynamics, a version of the model has been coupled to a free online simulator (“Behavioral Infectious Disease Simulator”, Struben 2020) that enables users to explore the impact of government and citizen responses, and examine how to alter the course of a pandemic. In the remainder I provide a short background of the COVID-19 pandemic, the responses, and of the existing relevant literature. I then perform a number of calibrated and stylized simulations. The paper concludes by discussing next steps.

## Background

### The virus

From December 31 2019 to January 3 2020 44 patients with pneumonia of unknown etiology were reported in China. On January 07 2020 a new type of coronavirus was isolated by the Chinese Ministry of Health (WHO, 2020b). Soon the Chinese Ministry of Health reported the cases’ exposure history to the Huanan Seafood Wholesale Market in Wuhan. The second week of January 2020 other countries identified confirmed cases related to traveling overseas including Japan, Thailand, South Korea. Reported cases went from over 150 thousand by mid-march to over 1.25 M on April 6th with over 65 thousand reported deaths in a total of 180 countries (Roser, Ritchie, and Ortiz-Ospina, 2020; Korean CDCs, 2020).

The virus causing the respiratory illness Coronavirus disease (COVID-19), “severe acute respiratory syndrome coronavirus 2” (SARS-CoV-2, earlier provisionally named “2019 novel coronavirus” (2019-nCoV)), is thought to spread from person to person through droplets and contacts when a person with the virus coughs or sneezes and by touching objects contaminated with the virus, then touching one’s eyes, nose or mouth. (Hereafter we solely use the acronym COVID-19, indicating both the virus and the disease.) Main symptoms include those associated with respiratory infections, ranging from mild to severe, such as fever, malaise, cough, shortness of breath and pneumonia. In addition, phlegm, sore throat, headache, hemoptysis, nausea, and diarrhea also appear. Elderly, immunocompromised patients, and patients with underlying medical comorbidities are most likely to be in critical condition or die from the virus. Early estimates suggest a crude case fatality rate for COVID-19 of about 1-2% (Shim et al., 2020; WHO, 2020b), much larger than the order of 0.1% for a moderate seasonal influenza. Yet, there is still much uncertainty about the true infection fatality rate (IFR) – of actual cases. Deriving this requires information about the actual number infected (the denominator), but this is challenging because the often limited testing capabilities and because endogenous factors such as hospital overload affect mortality risks (Ghaffarzadegan and Rahmandad, 2020). The IFR is particularly hard to detect, because of the large number of cases with mild and/or flu-like symptoms. For example, some studies suggest that about 80% of people with COVID-19 has mild (or no) symptoms, while 20% has severe symptoms, with about a third of those latter group becoming critically ill (ECDC, 2020).

The extent of an epidemic outbreak is affected by key virus transmission parameters. Estimating values of parameters such as infectious contacts and duration of infectivity is of critical interest to those seeking to impact this (Anderson et al., 2020). Compared to Influenza or Ebola transmission is rapid due to high infectivity. The duration of the infectious period for COVID-19 is estimated to be 5-10 days (Zou et al. 2020), after an incubation period of 2-14 (5.5 average) days (Li et al. 2020). The fundamental metric transmissibility of a virus, the basic reproduction number R_0_ – representing the number of people infected during once infectivity at the first infection – is estimated to be on the order of 2.4-3.3, quite higher than that for seasonal flu or Ebola (Chowell et al. 2004) but lower than for severe acute respiratory syndrome (SARS) (Lipsitch et al. 2003; Read et al. 2020; Walker et al. 2020). Estimates using this reproduction number suggest that globally, an unmitigated COVID-19 epidemic would lead to about 7.0 billion infections (Walker et al. 2020). Given the case fatality estimates, this could potentially result in 40 million deaths.

An unmitigated scenario, while important as reference, is unrealistic because transmission rates decline as governments and citizens will respond as reported cases and deaths accumulate, leading to reduced contacts. However, with such a high basic reproduction number outcomes must be seen in not only the total number of deaths, but also the peak load on the health systems and the risks of resurgence. Therefore, key policy questions are what set of responses and their timings help manage the outbreak path, in the short and longer run (Ferguson et al., 2020; Pueyo, 2020), and at what cost (Eichenbaum et al., 2020).

### Policy questions

Responses in Asia (South Korea, Hong Kong, Singapore, Mainland China, and, to some extent, Japan) show that active policy measures such as quarantine, social distancing, and isolation of infected populations can contain the epidemic (WHO, 2020b). While the outbreak has been contained within multiple countries through early government action and through social distancing measures taken by individuals, in many other countries this has not been the case.^2^ To illustrate, consider the outbreak and responses in three countries across three continents: South Korea, Italy (the first known epicenter in Europe), and United States (Figure 1). The three graphs show respectively cumulative reported cases (per million people pmp), cumulative tests performed (pmp), and metrics of relative social contacts, starting from the day of the first case reported to the WHO (December 31, 2019, time = 0 in the Figure) until May 12 2020 (time = 136).^3^ Across the three countries, the first reported case occurred all within one day (January 18-19 2020, Figure 1, top left). Initially, reported cases were much higher in South Korea, suggesting it had become an epicenter. However, the fate of the countries differed considerably during the following 90 days: Whereas in South Korea reported cases stabilized under 200 pmp by early March, in Italy by the end of March it was at 1700 cases pmp whereas in the United States, by the end of March, there were already 527 reported cases pmp. Case reporting however, is not independent from case testing. In South Korea it took 43 days to get from the first reported case to 2500 tests pmp, at which point there were 68 reported cases ppm. Italy reached 2500 tests pmp 15 days later with reported cases ppm at 291 ppm. The United States, reached this on March 27, 26 days later than South Korea, at which point there were 260 (steeply growing) cases ppm. With exceptions such as Iceland testing has lagged more in many other countries. In terms of social contact reduction, the data suggest that while South Korea was also relatively quick to respond here, Italy, once containment efforts began, responded more extensively to reduce social contacts. Notably, the United States were slower and less extensive in their response.

**Figure 1.**
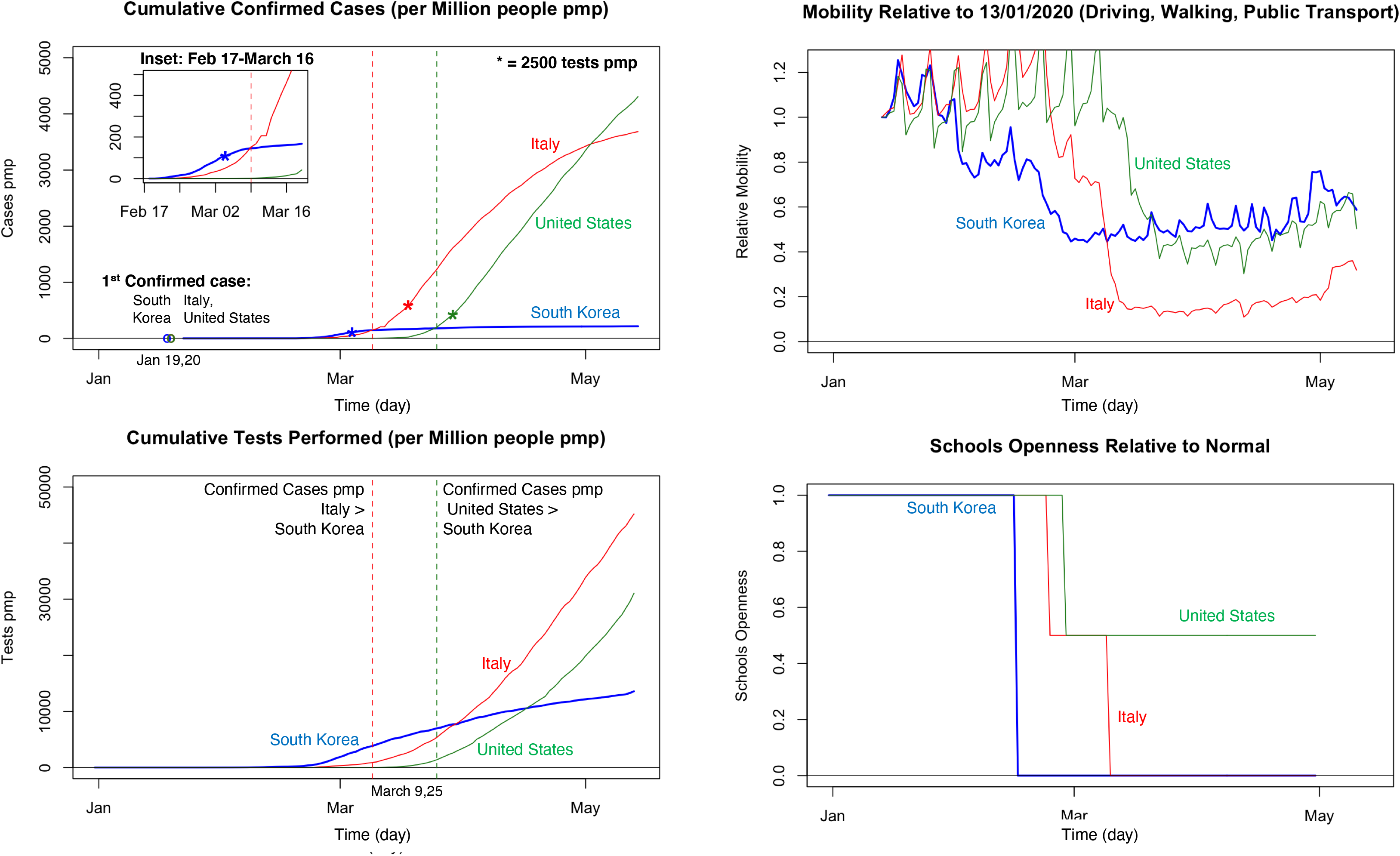
Data characterizing first-wave outbreaks and responses December 31 2019 - May 15 2020, for South Korea, Italy, and United States, showing reported cumulative cases and tests (per Million people, pmp), and social contact metrics. Data sources: Our World in Data (Roser et al. 2020); Mobility Trends Reports (Apple 2020); Unesco (2020).

South Korea’s approach reflects a more proactive approach beyond just testing. In particular South Korea has focused on early detection of persons at risk and so to identify and then isolate the virus (Korean CDCs, 2020). As part of this South Korea implemented a policy of early and widespread identification of suspected cases through targeted testing and isolation of “suspected cases” - family members and, through contact tracing, those that are thought to have been in close contact with positive cases. The quarantining and surveillance of positive and suspected cases involves follow up according to specific protocol and timing. Finally efforts are done to build capacity of local government, build systems of cooperation between affiliated organizations, and educate and raise public awareness among the community (Korean CDCs, 2020). In Europe confinement policies have been implemented but often much more slowly. Yet, they have focused more on confinement of the general population. Italy’s general lockdown commenced in the center and was gradually expanded to northern provinces (March 8). In the United States, despite urging from public health experts, by early April some states and counties have taken limited action, with beaches and restaurants still open (Axelrod, 2020). Indeed, social activity has taken much more time to slow in Europe and North America (Figure 1, bottom showing short term Air B&B lettings and Mobility trends for places like public transport hubs such as subway, bus, and train stations).

These contrasting outcomes and efforts highlight the importance of aggressive and extensive testing social distancing, often practiced in epidemic outbreaks (Jefferson et al. 2008). In line with this, during the first-wave COVID-19 outbreak delays in testing and social distancing may greatly augment deaths, directly from increased COVID infections, and indirectly through overburdening of the health system (Greenstone and Nigam 2020). Analyses of the merits of specific actions must also consider more targeted approaches such as contact tracking, suspect case isolation, and community surveillance (Fraser et al 2004; Wong et al. 2016). Further, they must also cover deconfinement and resurgence efforts. As countries have responded differently to the first wave and have begun making decisions towards deconfinement it has become clear that their decisions are often influenced by complex pressures across realms – reports of reduced load on critical care units, economic actors with calls for opening business, frustrated citizens subjected to long-confined societal members, neighboring countries with their own actions. Absent a thorough understanding of the dynamic interplays across virus transmission, citizen responses, and policy realms making an informed decision about altering - delaying, relaxing - interventions, individually or jointly, is impossible. A well-grounded dynamic integrative analysis is critical for developing such understanding.

## The Model

The computational policy model I outline here focuses on policy interventions for an infectious disease outbreak throughout the epidemic period, including deconfinement and resurgence (Figure 2, high level overview of the model). To capture virus transmission dynamics the core of the model forms an epidemiological compartmental SEIR structure commonly used by epidemiologists (Hethcote, 2000), with explicit times to symptom onset and infectiousness, that is consistent with earlier epidemiological studies (Figure 2 top left). The model is also sensitive to the socio-behavioral complexity of policies and citizen response with various ways to reduce social contacts of the general and infected population (Figure 2, bottom left). Policy makers make choices about building testing and capacity (that allows identifying positive cases) and testing strategies that allow targeted approaches that identify cases within clusters and allow for surveillance of positive and suspect cases (Figure 2, top right). To model traces the population in its different stages (Figure 2, note on the bottom right).

Besides metrics such as reported cases, actual cases, and deaths, socio-economic pressures such as hospitalizations and aggregate reduction in social contacts. To study the impact of policy interventions and citizen responses and the variations of those policies, of citizen responses, and of the virus dynamics, across geographic and political regions the model is disaggregated into N segments. These segments can be used to represent geographical regions such as continents, countries, or provinces for example (as long they are sufficiently large so that micro-level variation in individual contacts are less important), but also to represent socio-demographic variation (older versus younger populations; vulnerable versus less vulnerable groups) within a single geographic region. Finally, a number of assumptions can be altered that affect the evolution of endogenous contacts and case testing and reporting, and through those, virus transmission dynamics (Figure 2 bottom right).

In what follows I highlight the key model concepts, structures, and variables. Likewise, the accompanying figures show simplified representations of the model sections. A technical appendix lists all model equations, using the same sections and sequencing as below and provides additional visuals (Appendix A.I).^4^

**Figure 2.**
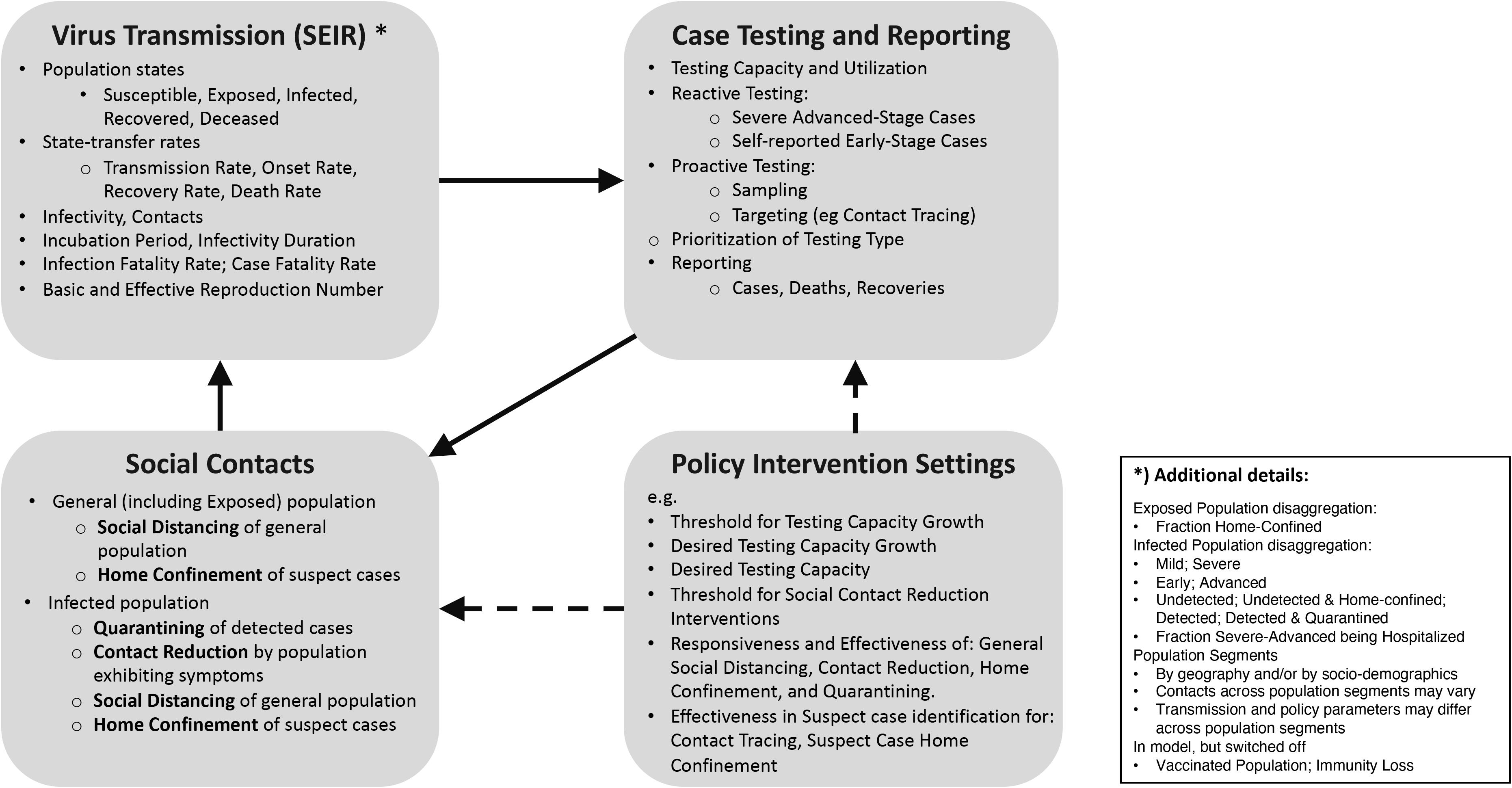
High level model overview.

### Virus transmission

In the SEIR structure (Figure 3) the infectious population transmits the virus to susceptible population within demographic segment *d, S_d_*, through infectious contacts *ic_d_* at transmission rate *tr_d_ = ic_d_* · *S_d_*. Infectious contacts may come from the population in the infected state (typically, but not necessarily associated with the onset of symptoms), within demographic segment *d’ I_d_*, as well as from the exposed population *E_d′_*. Infectious contacts depend on susceptible population being in contact with those populations, with respective contact rates 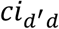 and 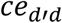 and on infectivity - the probability of infection given contact between a susceptible and an infectious person. Then, the virus transmission (in simplified form)^5^ is given by:

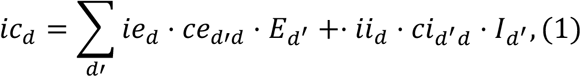

Infectivity of the infected population, *ii_d_*, tends to be considerably higher than that of the exposed population, *ie_d_*. However, viral load measures suggest that in the case of the COVID-19 infectivity commences before the onset of first symptoms (Ferguson et al. 2020; Pan et al. 2020; Zou et al. 2020;) – thus during the exposed period. Further, infectivity may to vary across segments because regional climate (temperature/humidity) affects transmission (Xu et al. 2020; Kissler et al. 2020a). *ce_d_*_′_*_d_* and *ci_d_*_′_*_d_* equal within-segment contacts *ce_d_* and *ci_d_*, adjusted for relative cross-segment contacts *fc_d_*_′_*_d_*. For example for the infected population: *ci_d_*_′_*_d_ = fc_d_*_′_*_d_* · *ci_d_*_′_. (We discuss the within within-segment contacts below.)

After transmission, the population remains in the exposed state during a latent or incubation period *λ* followed by the onset of the virus infection (Figure 3, Onset Rate from Exposed to Infected Population). At this point they (may) begin to show symptoms. Below we outline in more detail the more disaggregated formulation of the infected state, including those parameters marked in Figure 3 with “*”. Finally, depending on the (true) infection fatality rate (IFR), those in the infected state either recover or die (Figure 3, right).

**Figure 3.**
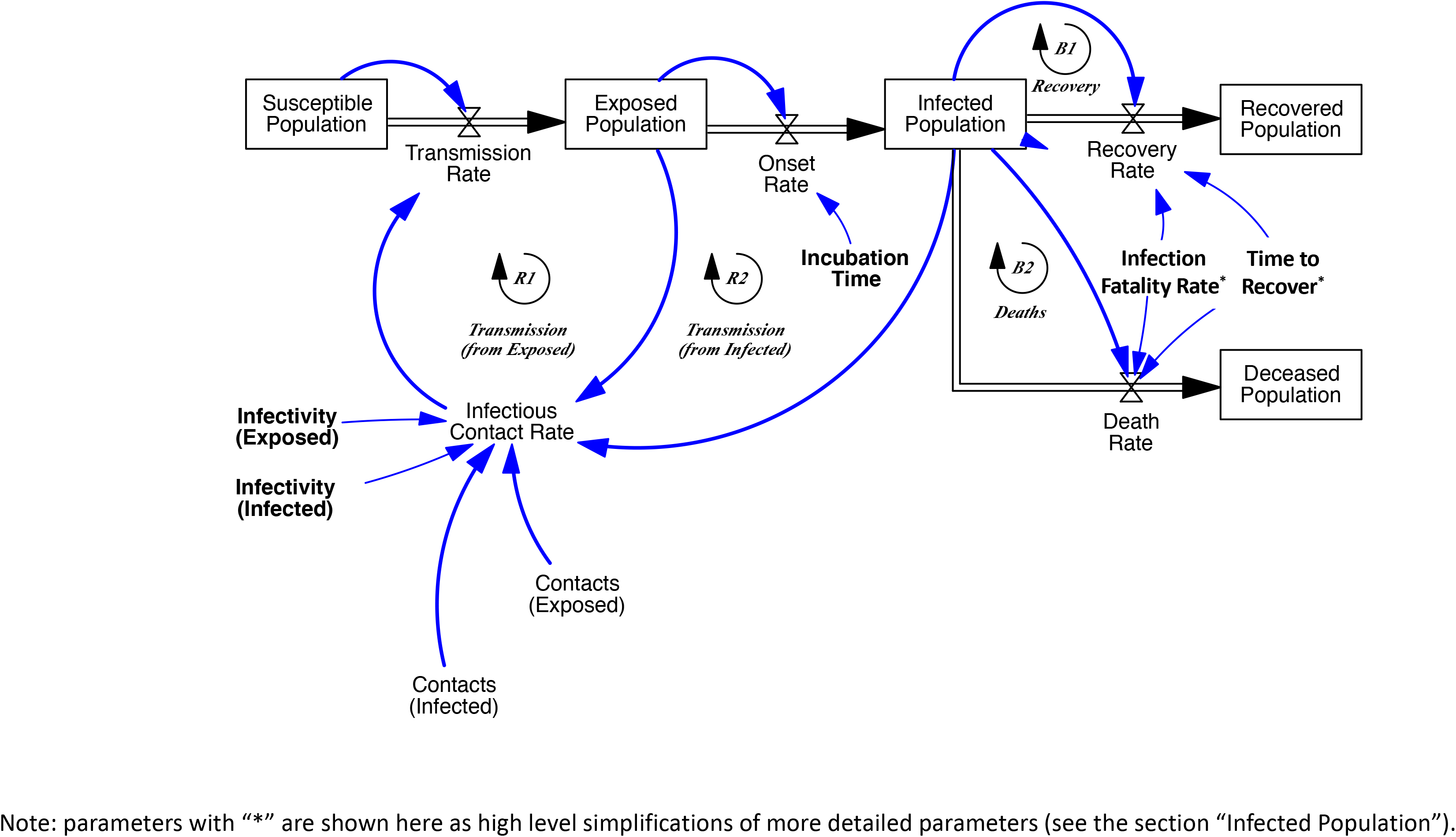
Virus transmission structure (simplified representation). Note: parameters with “*” are shown here as high level simplifications of more detailed parameters (see the section “Infected Population”).

### Infected population

The structure of the infected population is further disaggregated, most importantly to capture the role of the large variation in symptoms across the populations and, because of that, low detectability for part of the infected population (Figure 4). Symptoms may vary considerably across those infected, with only a small fraction of those infected having severe symptoms (ECDC, 2020). In the model the fraction of severe cases *fs_d_* is defined as the fraction of all cases requiring some form of critical care. A share *fh*s of those with severe symptoms actually gets hospitalized, once they progress from early to advanced stage, after time to reach the advanced stage *τ_a_* (Figure 4, top). Next, these severe cases either recover or die after a time to recover *τ_rs_*, with the severe-case infection fatality rate *frs* being the actual (not reported) fraction of fatality fraction of the severe cases. Those with mild symptoms (Figure 4, bottom) - including a large share being fully asymptomatic - follows the same two-stage structure. However, none of those are hospitalized or die, so all recover after the recovery time for the mild population *τ_rm_*. (Thus, the IFR equals *ifr_d_ = ifrs · fs_d_*).

**Figure 4.**
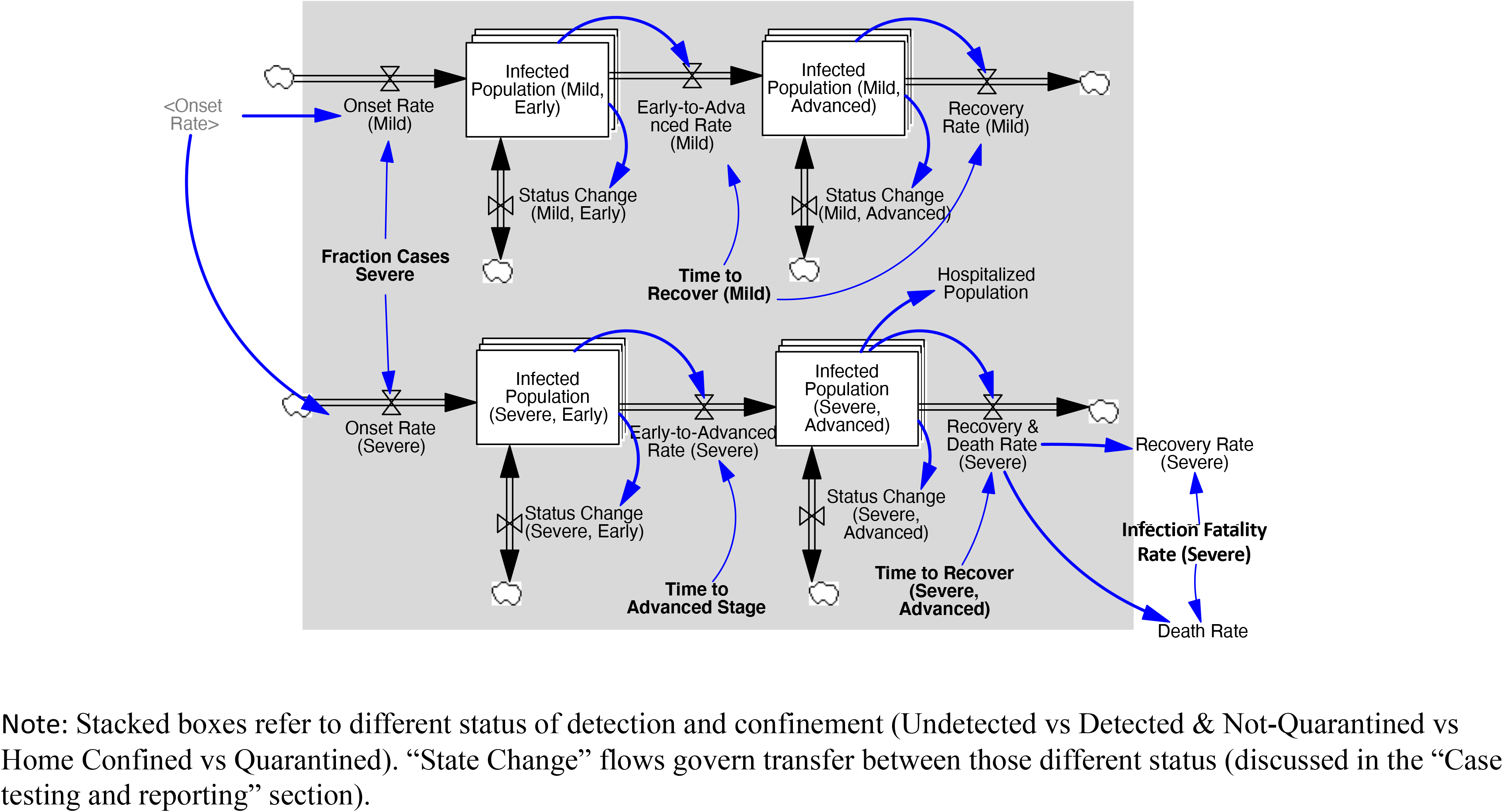
Infected population (detailed representation). Note: Stacked boxes refer to different status of detection and confinement (Undetected vs Detected & Not-Quarantined vs Home Confined vs Quarantined). “State Change” flows govern transfer between those different status (discussed in the “Case testing and reporting” section).

### Social contacts

Virus transmission depends not only on exogenous virus-related transmission parameters but also on citizens and policymakers responding to an outbreak and taking measures to protect themselves or to curtail this outbreak. Further those with symptoms may reduce contacts because they stay at home feeling seek, irrespective of concerns about the virus outbreak. As the population adjusts social contacts, infectious contacts and transmission rates change too.

In the model social contacts of the infected population as well as of the general (and thus exposed) population reduce in response to the severity of a perceived outbreak in a number of different ways (Figure 5). The infected population may reduce contacts as: i) positive tested population (detected cases) - at hospitals or elsewhere – are being quarantined (***quarantining***); ii) undetected cases exhibiting symptoms reduce their contacts voluntarily or urged by governments, beyond what they would otherwise do because of sickness (***contact reduction***); iii) undetected cases reduce contacts either voluntarily or being urged by governments (***social distancing***). General contact reduction may range from increasing washing hands, to prohibitions in gathering in groups, travel restrictions, to school and non-essential office closings, mask waring, etc.; iv) undetected cases are being home-confined because they have been associated with detected cases through targeted isolation efforts (***home confinement***). The exposed population may reduce contacts because: i) undetected cases reduce contacts either voluntarily or being urged by governments (***social distancing***); ii) undetected cases are being home-confined because they have been associated with detected cases through targeted isolation efforts (***home confinement***).

**Figure 5.**
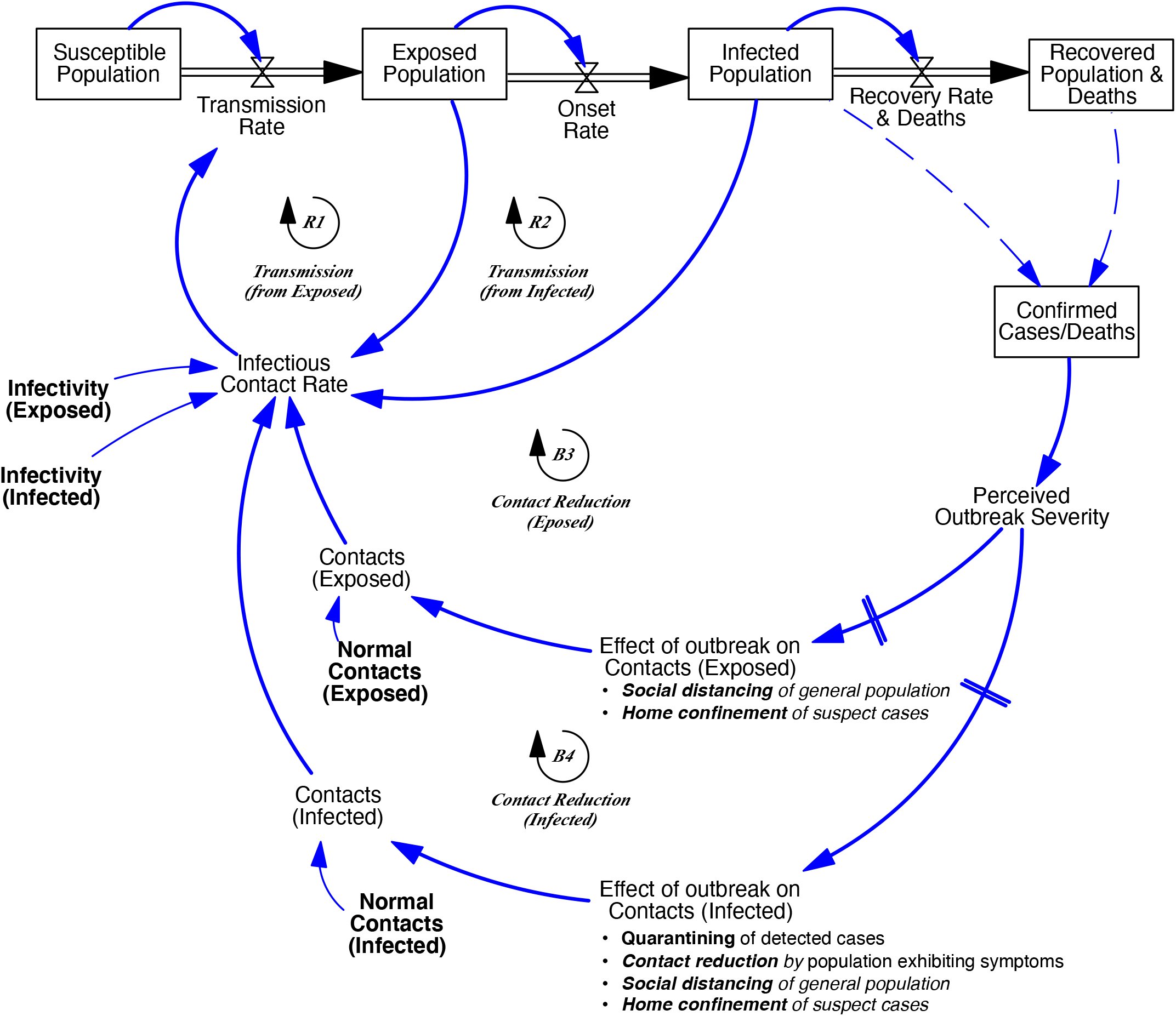
Social contact reduction in response to perceived outbreak severity (simplified representation).

Social contacts, for both the exposed and infected population, *ce_d_*, and *ci_d_*, form a weighted sum across population groups undergoing one or more of the contact reductions, for thus, for the infected population, *ci_d_ =*Σ*_q_ci_dx_*, with *x* ∊*{u undetected, d detected, q quarantined}*. We model this through multiplicative contact reducing effects based on a pre-outbreak reference contact rate *c_norm_*. Thus, for example, indicated contacts of the undetected infected population *ci_du_*, with home-confinement effect *icf_d_* and with general social distancing effect *isd_d_*, are:

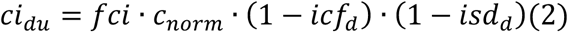

where *fci* is the relative contact rate of infected population (compared to the normal contact rate of the general population).

Contact reductions for those within their state adjust to indicated levels of contact reduction over adjustment time *τ_c_*. For example, for social distancing for the infected population, *isd_d_* adjusts to the level indicated by general social distancing 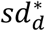, thus 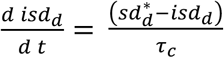. Those quarantined and home-confined adjust contacts as they are being transferred to their new state. Finally, those newly infected adjust their behavior to that indicated by the infected population over time. This adjustment of contacts of newly infected population is captured through a co-flow structure (Sterman 2000).

For the formulation of the social contact adjustments through each of the above effects we cannot rely on a prior body of literature and we therefore develop this here based on behavioral grounds. Such a formulation must include three factors: First, citizens and policy makers adjust contacts based on the perceived severity of the outbreak. Second, the extent of adjustment depending on how sensitive they are to increasing levels of the perceived outbreak. For example, achieving citizen responsiveness requires clear government communication and media and awareness campaigns. Third, there are limits to how much one is able or willing to reduce contacts, irrespective of the extend of the perceived outbreak. This limit may come from implementation challenges (quarantining of patients may still lead to health worker infection), practical limits (those being home-confined still need to go out to buy groceries), or, simply, because not everybody complies and enforceability is limited. To capture sensitivity to the outbreak, we follow a general formulation for all contact reduction variables with as input the perceived outbreak level *o_d_* relative to some reference outbreak level *o_ref,d_*. We define the reference as the level at which any response to the breakout commences. Continuing the illustration of the social distancing effect *sd_d_*, with sensitivity *β_sd_,_d_* and maximum contact reduction *sd_max,d_*the formulation is:

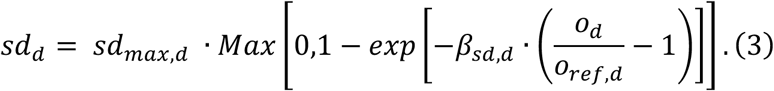

This formulation implies that below reference outbreak level *o_ref,d_* there is no social distancing effect. At *o_ref,d_* the marginal responsiveness is maximal and subsequently decreases till it asymptotically reaches *sd_max,d_*. A greater *β_sd,d_* implies higher marginal individual responsiveness or larger populations.

As policymakers and citizens alter their behavior in response to the severity of the perceived outbreak and so affect virus transmission rates they respond to reported (not actual) data about positive tests and deaths. Media and experts report different metrics about the virus but reported absolute cases and deaths tend to dominate the media and affect population behaviour (Xiao et al. 2015). The perceived outbreak level *o_d_* is a simple weighted function of the reported deaths *RD_d_* and reported cumulative cases *RC_d_*, *o_d_ = w_d_RD_d_ +* (1 *− w_d_*)*RC_d_* ^6^

### Case testing and reporting

Case testing follows two main approaches: reactive and proactive. First, reactive testing is driven by the symptoms occurring under the currently undetected infected population *I_iu_* (omitting demographic index *d*) within any of the states *i* either being mild *m* or severe *s* cases in the early *e* or advanced *a*, thus *i* ∊ [*me, ma, se, sa*} (Figure 4). Reactive testing occurs when the undetected population either self-reports their symptoms or is hospitalized with symptoms. The reactive testing process is identical across all states *i* (Appendix Figure A. 1 visualizes the testing process for the severe, advanced infected population *I_sa,u_*). Correctly identified positive tests *tp_i_* (hence, ignoring false positives) equal the fraction of actual cases tested *t_i_* times the case detection fraction *fd*, and the fraction of actual tests being positive *fp_i_* (one minus the false negatives): *tp_i_ = fd · fp_i_ · t_i_*. The actual testing rate equals the desired testing rate constrained by effective testing capacity available for i, *tce_i_*.

Hence, 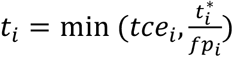. Desired testing *t\** results from a fraction of the population *I_iu_* reporting symptoms of which a maximum fraction and is deemed acceptable for testing, together captured by 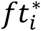. (The Appendix (section A.I.3) details aggregation from and allocation of testing capacity across the different infectious states *i*). With the infectious cohort time *τ_i_*, the indicated test rate for infectious population in state *i* is defined as 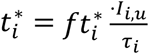.

Second, proactively, experts can perform field tests, provided available capacity. There are two types of field tests: “sampling” and “targeted” testing, indicated by index *f ∊ {sp, ta}*. Targeted testing may involve methods such as contact tracing testing and occurs within a small targeted sample of the population with relatively high likelihood of positive case detection. However, this approach requires efforts identifying potential positive cases. The process (highlighted in Appendix A.I.3, Figures A.2) involves first, positive testing, with time to identify and test potential case *τ_t_*, increasing with the number of tests performed within the effective population size (or catchment area) *N_f_*. However, the marginal value decreases in tests *t_f_* performed, with that of the first being equal to the effective density 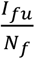. The solution for this problem is:

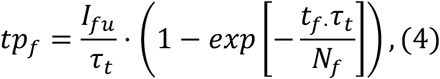

where, as with reactive testing, proactive testing is constrained by the capacity, hence 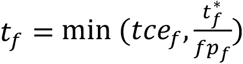*. N_f_* is the effective pool within which to search. Because neither search type is close to random, *N_f_* can be considerably smaller than the size of the population group within which the search if performed. We find by the ratio of size of the undiscovered pool *I_fu_* and the actual likelihood of finding a subjective case *p_f_*. This 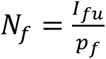 A clustering parameter *κ_f_* tunes the likelihood of a positive tests (with random probability being the base likelihood). The adjusted likelihood of a positive test corrects for the maximum fraction of potential cases accepted for testing 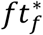 and for detection fraction *fd:*

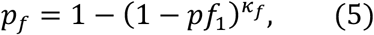

where *pf*_1_ *= fd · ft_f_ · pr*, and *pr* is the likelihood of a single undetected infected person 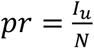. The targeted search effectiveness parameter *κ_f_* captures the effectiveness of proactive testing by indicating how efficiently “detectable cases “ (those that have been infected by others) are actually identified and tested. This formulation implies that at small probabilities (*pf′* ≪ 1)*, κ_f_* approximately acts to linearly increase the probability of success (*p_f_ ≈ κf · pf*_1_) and thus proportionally decreases the effective search space 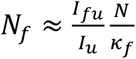. (The formulation of Eq. (5) assures robustness for larger values of *κ_f_ · p_f_*.)

To illustrate, consider a population of *N =* 16*M* and *I_u_ =* 16k undetected cases in total (ie 0.1% of the population, so *pf*_1_ = 0.1%, assuming for simplicity *fd · ft_f_ =* 1). Let *I_u_* =1000 of those cases exist within some known cluster of high infections, and *t_f·_* =50 tests being performed within those known zones, with search time *τ_t_*=1 day. Then, a targeted search effectiveness *κ_f_ =* 1 (random search) would give about 0.05 expected positive tests. Then, using the approximation 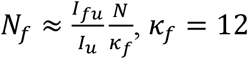 implies that 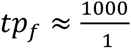. 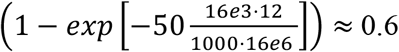 positive tests, while *κ_f_* = 24 gives ≈ 1.2 positive tests, and *κ_f_ =* 480 gives about 23 out of 50 tests being positive. (When using the model Equation 5 for *p_f_*, *κ_f_ =* 480 gives about 19 out of 50 tests positive - one can observe the diminishing returns in *κ_f_*.) The value of targeted search effectiveness *κ_f_* depends much on conditions affected by social structures and mobility of the population as well as on the capabilities of experts in tracing actual positive cases within hot-zones given such structures and mobility.

In addition, for contact tracing to be effective a sufficient large amount of amount of positively tested cases need to be traced. For sampling, simply *I_sp,u_ = I_u_*. For targeted testing the pool of detectable infected population *I_ta,u_* depends on active work done to trace the tested population and identify potential suspect cases (Appendix A.I.3, Figures A.3). Thus, *I_ta,u_*builds with the detection of new cases *tp*, depending on the ability to identify associated cases through targeted testing *fata* as well as on the effective cases each infects, given by the reproduction number *R*. Thus, new detectable cases build with 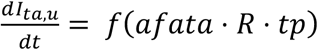. The actual change rate is contained with the size of the pool of undetected cases and has outflow proportional to the loss rate of undetected cases. (Appendix Figure A.3 shows the relations.)

Testing is capacity constrained by availability and utilization of testing kits *TK* (omitting index *d*). (See Appendix Figure A.4.) Initially a fixed number of kits *TK*_0_ is available. When cumulative hospital visits associated with the virus symptoms exceed threshold level *CH^*^*, the building of additional testing kits begins, at growth rate *g*. The production of test kits continues until a desired level of testing capacity is achieved (cases/day/million people). Utilization of test kits is a function of the growth rate of reported active cases: If reported active cases decline (increase), utilization reduces (increases). Finally, testing capacity is distributed across testing strategies, according to the following priority rule: i) reactive – severe-advanced population; ii) reactive - self-reported symptom-based of early stage severe and mild cases; iii) proactive - field testing (targeted); iv) proactive - sampling.^7^

### Quarantining and home confinement

Those who test positive are quarantined at quarantined fraction *fq_i_*, and thus move at rate *fq_i_ · tp_i_* from undetected infectious *I_iu_* to quarantined infectious state *I_iq_*, while the remainder (1 − *fq_i_*) *· tp_i_* moves to the detected (but not quarantined) state *I_id_*. Home confinement of potential suspected populations (exposed and undiscovered infectious) builds from the stock of detectable cases for positive tests through contact tracing. The stock of detectable cases accumulates in the same way as for field testing. The rate of accumulation is enabled by an ability to identify associated cases for home confinement, *fahc*. (For more details see Appendix Figure A.5.) The actual home confinement rate depends on the fraction of potential cases being home confined *fhc_d_*. This fraction builds as a function of perceived outbreak in the same way as social distancing (Equation 3), and depends on the reference outbreak level *o_ref,d_*, the maximum home confinement fraction *crh_max_*, and sensitivity parameter *β_x_*.

## Analysis

Five logically ordered experiments illustrate the value and flexibility of the model for different policy analyses. The first experiment involves the construction and analysis of a Baseline run that builds, through calibration, on the ongoing COVID-19 outbreak using a cross-sectional dataset involving 6 countries. In the second experiment we perform a sensitivity analysis of this Baseline case, centered on policy and citizen responses to the outbreak within one of the countries. In the next two experiments we perform an analysis, within hypothetical regions, about managing deconfinement and virus resurgence. In the final experiment, we explore the relative effect of the virus on vulnerable populations, by varying the symptom severity across population segments.

### Experiment 1 – Baseline calibrated to the ongoing COVID-19 outbreak

To perform a cross-sectional calibration of the model I constructed a country-level dataset with diverse outbreak data including reported daily new cases, recovery rates, death rates, and testing rates, as well as a metric for social contact rates. The dataset covers a large number of countries from December 31 2019 until May 15 2020.^8^ Daily reported new cases and deaths and testing data were retrieved from Ourworldindata.org (Roser et al. 2020). The metric for social contacts is a composite of data on intensity of walking, driving, and public transport use and of data on school closings (See Figure 1 for details and sources). To obtain the social contact data I multiplied a reference normal contact rate value, constant across countries, with the population size. The full dataset includes over 200 countries and can be customized to select a subset of countries and to aggregate across a subset of countries. The dataset can also easily be updated as new data become available.

I then strategically selected six countries for in-depth analysis, aiming for variation in outbreak dynamics and interventions and selecting for the presence of reliable reporting data for calibration. The final set includes the three countries already highlighted in the background section plus three others, combining to: South Korea, Germany, Italy, France, Sweden, and the United States (S-K, GER, ITA, FRA, SWE, and US) (Table 1). Within this set, after South Korea, Germany has the lowest cumulative reported cases per capita (though considerably higher than for South Korea). Germany responded relatively quickly, in particular through early testing buildup and isolation of suspect cases. France has, like Italy and the United States, a relatively high number of cumulative reported cases. While France was relatively slow to expand testing capacity, it did impose very strict and extensive confinement rules as of March 17. In all countries but Sweden active cases have stabilized or are (for now) on the decline. On May 15 2020, active cases of Sweden are still growing (Table 2). Sweden followed a deliberately moderate confinement approach and was also slow to build up testing (Table 2, maximum relative social contacts).

I then designed a single cross-sectional calibration of the model for the 6 countries combined using Log Likelihood-based estimation of parameters and of their unitary confidence intervals (conform Ghaffarzadegan and Rahmandad, 2020; Keith et al. 2017; Struben et al. 2015). To limit the large set of potential variables to be estimated, I set virus-transmission parameters for which the existing empirical literature on COVID-19 has already produced reliable estimates (e.g. incubation time). I focused the final calibration on a subset of 22 parameters (remaining parameters were set heuristically or set using initial partial model tests). The cross-sectional, joint calibration performed here is preferable above individual calibrations because several of the parameters are expected to be to a great degree independent of country. In such cases the joint calibration can give better insights into some the mechanisms at work and demonstrate the robustness of the model behavior. In this case, of the 22 estimated parameters 11 were estimated across countries (Table 2) and 11 within countries (Table 3). (Their estimated values are discussed below.)

Figure 6 reports the calibrated outbreak data, being new reported cases, death rates, testing rates, and total social contact rates (omitting new reported recoveries), and their simulated values across the 6 countries. Overall, the calibrated simulated data replicate a smoothed path of the noisy data.

More important, the parameter estimates generally fall within plausible ranges (See Table 2 for cross-country virus transmission and clinical parameter estimates and Table 3 for country-specific parameter estimates). To illustrate, the relative normal contacts of infected population are considerably lower than the normal contact rate of the general population (*fci* = 0.164). Because of adjustment delays to symptom-based behavior this effectively results implies, controlling for infectivity, about a 67% reduction in social contacts (absent interventions). Estimation of infectivity of the infected population *ii*=0.635, together with those of other virus transmission parameters allows calculating infectious contacts and initial growth rate of the outbreak, as measured by the basic reproductive number R_0_. The basic reproductive number R_0_ is defined as the average number of secondary infections produced when one infected individual is introduced into a host population where everyone is susceptible (Dietz, 1975). Thus, a value of R_0_ > 1 implies active cases grow and that an epidemic can get started. The parameter estimates of the basic reproductive number of around R0=2.39 are close to other estimates (Read et al. 2020). On average, 4.2% of the cases is estimated to be severe, within the definition of the model, implying that about 5% of all cases are hospitalized. Based on the fraction severe cases and the infection mortality rate for the severe cases *ifrs* = 0.187, the actual estimated infection mortality rates (*ifr = ifrs · fs*) are estimated to between 0.67% (Germany) and 1.2% (South Korea), with 0.78*%* for the Unite States and 1.1% for France. This is roughly consistent with values other studies (eg Shim et al. 2020; Ferguson et al. (2020; WHO 2020b) estimating values between 1% to 1.5%. Those values likely overestimate the IFR due too the large number of cases exhibiting mild or no symptoms.The virus transmission parameter time to recover for the infected population with severe symptoms is estimated to be above 25 days. This value is consistent includes full recovery (or death) and is consistent with other findings (eg Ferguson et al. 2020). Note that the value exceeds the average duration of infectivity.

Estimates of contact reduction efforts highlight the considerable variation across the countries. Initial responsiveness to the outbreak across countries as indicated by (Table 2, Reference Outbreak Level, *o_ref,d_*) suggests higher initial responsiveness by South Korea and Sweden and lagged initial responses by Italy, France, and United States. The combination of a lower (higher) outbreak level and higher (lower) sensitivity parameters for a small country like Sweden (United States) is also consistent with population size effects giving rise to heterogeneity. Maximum contact reduction efforts varied considerably between the countries, and different from the initial responsiveness, with general population social distancing (*sd_max,d_*) reducing social contacts of the general population for five countries to about 49% - 88% compared to normal, with the largest effects in France (and Italy) and the smallest in Sweden and Germany (Table 2, maximum contact reduction fraction (general)). The estimates suggest however that Germany however, should be viewed differently from that of Sweden in that it developed, like South Korea, targeted home-confinement ability, as can also be seen from the parameter Relative Efforts Needed for Full Home Confinement Ability *efahc_d_*, with a value close to 0. In terms of other targeted approaches, estimates suggest that South Korea deployed targeted contact tracing aggressively. This is also reflected in the parameter “Relative Targeted Testing Needed for Full Targeted Testing Ability” *efata_d_*, also being close to 0. This parameter should be seen in combination with total testing capacity being available for such targeted testing, and indicates that South Korea was fully effective in contract tracing from early on in the process.

**Table 1.**
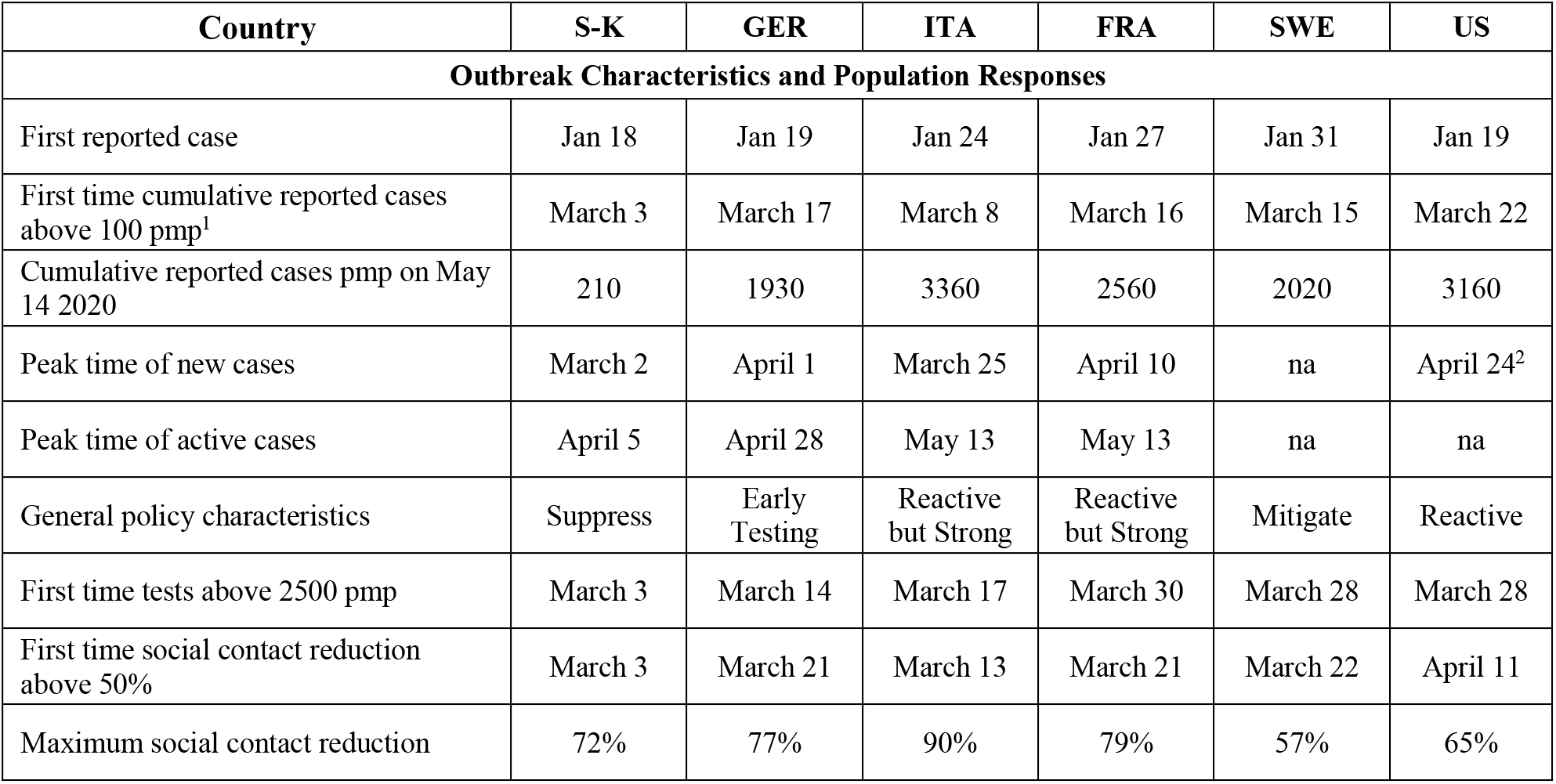
Country-Specific Outbreak Characteristics

| Country | S-K | GER | ITA | FRA | SWE | US |
| --- | --- | --- | --- | --- | --- | --- |
| Outbreak Characteristics and Population Responses |  |  |  |  |  |  |
| First reported case | Jan 18 | Jan 19 | Jan 24 | Jan 27 | Jan 31 | Jan 19 |
| First time cumulative reported cases above 100 pmp<sup>1</sup> | March 3 | March 17 | March 8 | March 16 | March 15 | March 22 |
| Cumulative reported cases pmp on May 14 2020 | 210 | 1930 | 3360 | 2560 | 2020 | 3160 |
| Peak time of new cases | March 2 | April 1 | March 25 | April 10 | na | April 24<sup>2</sup> |
| Peak time of active cases | April 5 | April 28 | May 13 | May 13 | na | na |
| General policy characteristics | Suppress | Early Testing | Reactive but Strong | Reactive but Strong | Mitigate | Reactive |
| First time tests above 2500 pmp | March 3 | March 14 | March 17 | March 30 | March 28 | March 28 |
| First time social contact reduction above 50% | March 3 | March 21 | March 13 | March 21 | March 22 | April 11 |
| Maximum social contact reduction | 72% | 77% | 90% | 79% | 57% | 65% |

**Table 2.**
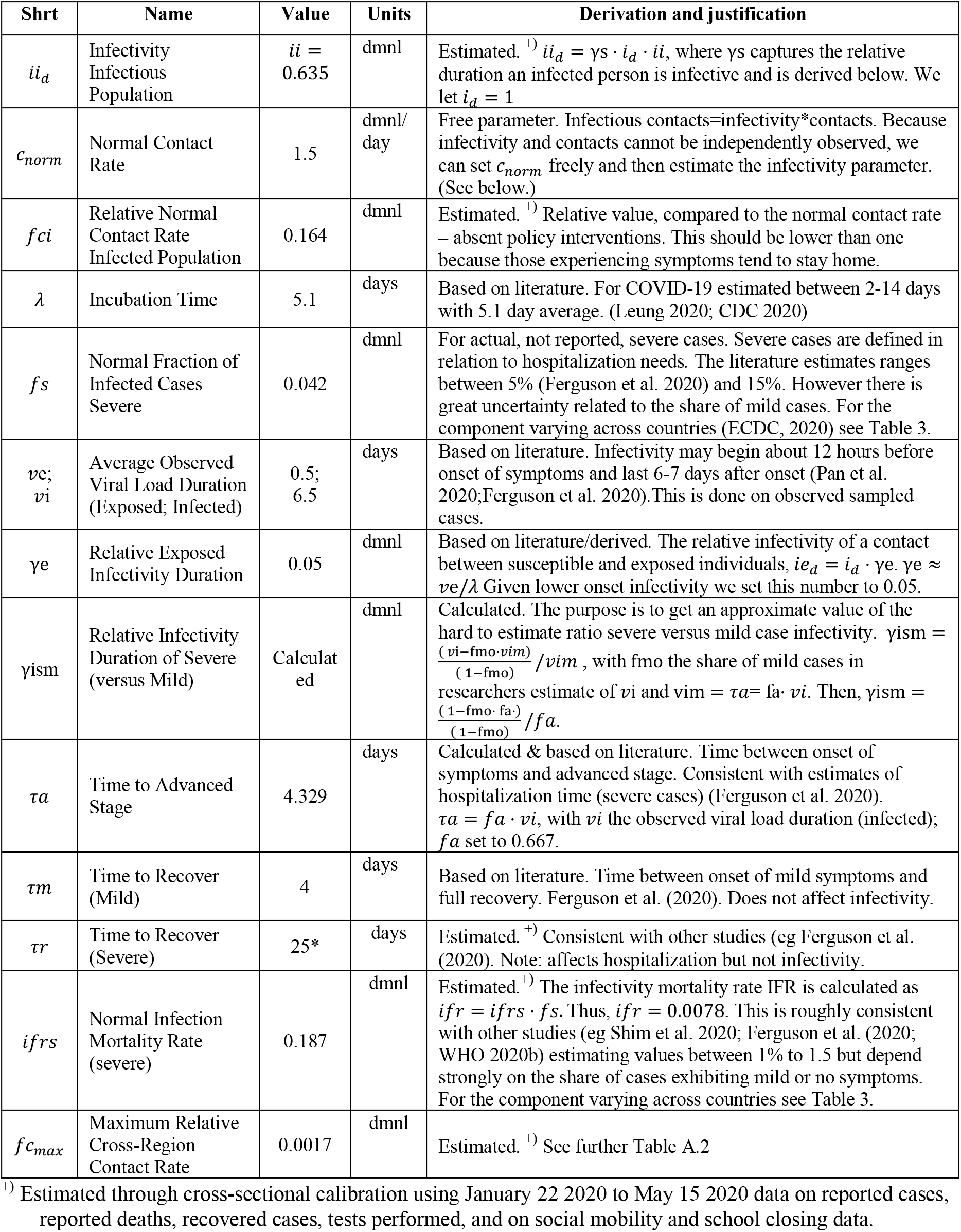
Estimated and otherwise derived values of virus transmission parameters fixed across countries. Estimated parameter values shown here are rounded for visual purposes.

**Figure 6.**
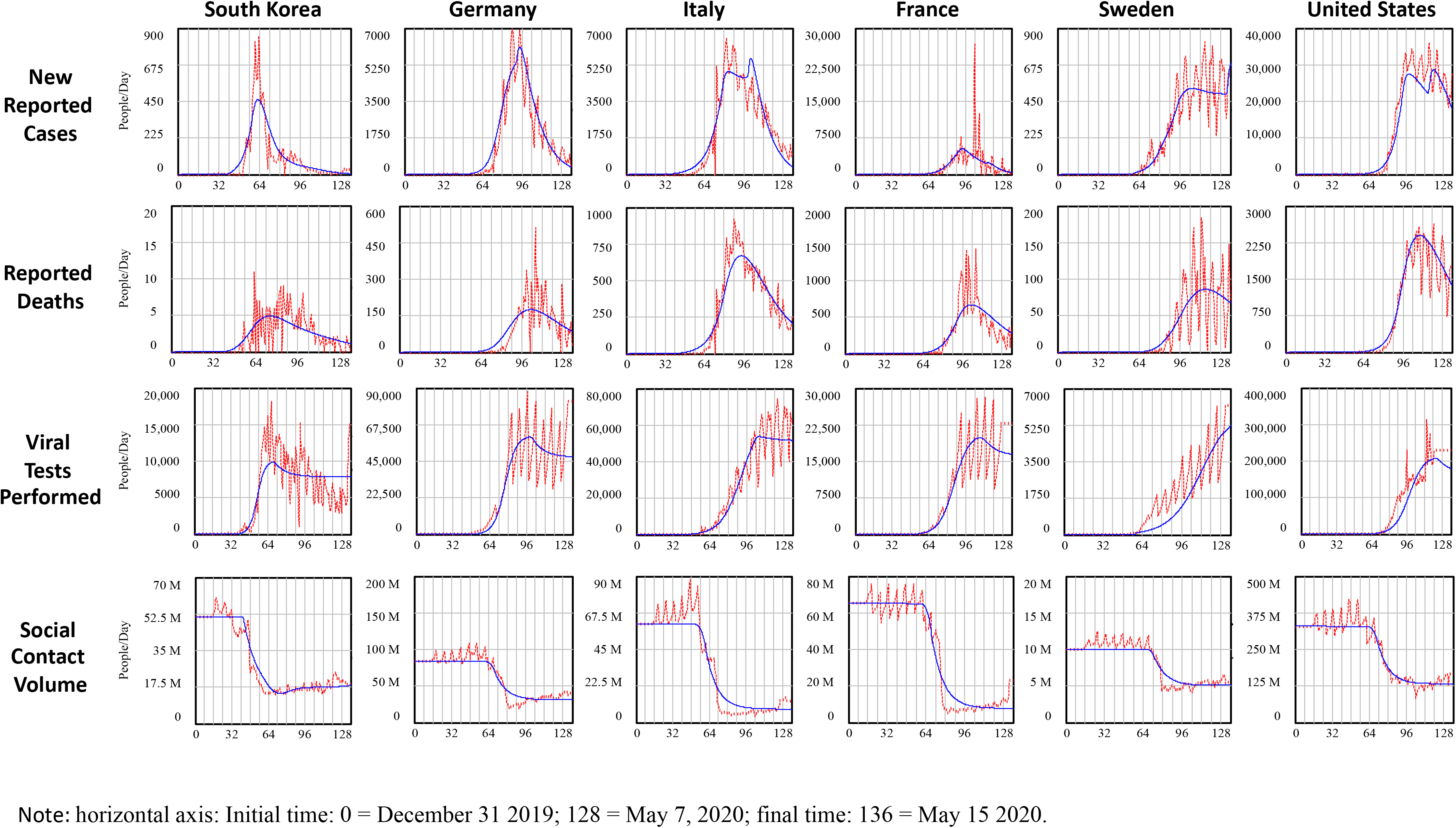
Calibrated simulation and data by country Note: horizontal axis: Initial time: 0 = December 31 2019; 128 = May 7, 2020; final time: 136 = May 15 2020.

Figure 7 shows more detailed results, including an out of sample period for a counterfactual scenario in which social distancing policies and behaviour remain in place as of May 15 2020.^9^ The calibration against data runs from time t= 1-136 (29 December 2019 - May 15 2020, shaded areas). The out of sample results run until t=250 (Sept 05 2020). The left panel depicts, for a subset of countries (South Korea, United States, Sweden), simulated reported and actual cases, as well as shows the actual data on reported cases for the relevant period (t=136). The data show very large variation in both reported and actual cases across countries. Moreover, their ratios vary considerably. In September 5 in South-Korea simulated actual cases are about cases are about 0.6 per thousand, while this is respectively 93 and 94 per thousand for Sweden and the United States. One can also infer that in South-Korea simulated reported cases are about 37.7% of simulated actual cases (thus suggesting that a little under 38% of all cases have been detected, this value is much lower in other countries. For the United States and Sweden these values are much lower, respectively 4.6% and 6.0%. The simulation also shows that whereas South Korea and also the United States have close to halted growth in cumulative cases, in Sweden, under a continuing social distancing policy cases likely will increase considerably increase. Not shown, Germany is at 23 per thousand and France and Italy are just above 100. Underlying these differences is the large number of mild (including asymptomatic) cases and the testing capacity constraints during the transmission growth phase there is a large gap between actual and reported data. However, there is more to it. The remaining graphs in Figure 7 provides additional details from the baseline simulation about the underlying dynamics driving these outcomes.

Figure 7 (centre, top and middle) shows simulated tests performed within two comparable easily countries, France and Germany, differentiated by three types: i) reactive testing of advanced-stage population exhibiting symptoms (all severe cases, Figure 4, bottom right stock). Most of those cases involve tests performed upon hospital admission (tests with high priority); ii) reactive testing of early-stage populations, once they exhibit symptoms (mostly involving severe cases, Figure 4, left stocks); iii) proactive testing through field tests including through targeted approaches such as contact tracing. Simulated testing for France (centre, top), shows a that initially most tests involved severe advanced-stage cases. By contrast, within Germany early on, many of the tests performed involved also earlier stage case testing. While these differences can be partly explained by differences in the early ramp-up of the testing capacity (compare the total tests performed of Germany and France), the dynamics are more complex than that. A second part of the explanation for the differences in types of testing is the result of differences in deliberate choices in testing approaches. To highlight this further, consider also the case of South Korea. The baseline simulation highlights that South Korea was able to perform pro-active testing even earlier than Germany. Figure 7 (center, bottom), illustrates this, showing South Korea’s high case detection fraction of not only severe but also mild cases, in contrast to those for the United States and Italy. The simulation shows that South Korea’s developed contact tracing ability (Low Relative Targeted Testing Needed for Full Targeted Testing Ability *efata_d_*) has had a considerable impact on the positive tests. However, the testing approaches are to great degree endogenous. Once a country falls behind in testing, a good part of testing capacity must be deployed to test severe cases. In this way, however, early stage cases remain undiscovered, reducing the opportunity to identify and isolate knowable cases. In turn, those infected will maintain social contacts for longer durations, leading to more transmissions. This then, leads to more cases in the long run and, through that more testing capacity constraints. In the simulation, as in the real world, during the outbreak stage South Korea had a lower positive testing rate than other countries had during the outbreak stage. This is part because of the higher relative testing capacity, compared to other countries. However, because of their approach a greater share of their tests involves mild cases. With positive testing rates for the mild cases being much lower than those for severe cases, during an outbreak, with constrained testing, it is very difficult to move upstream towards testing earlier-stage and milder cases. Therefore, countries with testing constraints risk being pushed further and further towards reactive testing. Absent a capacity to identify positive cases in the first place, one can certainly not find others through targeted approaches. That is, a positive feedback acts to move efforts towards downstream-reactive testing - away from proactively identifying, testing, and isolating upstream exposed and symptomatic populations: once testing capacity falls behind, most cases are identified in the hospital, or through severe-symptoms in the late stage. (Figure A7 shows a causal loop diagram highlighting these dynamics in further detail). Effective targeted approaches thus require early testing buildup.

The top right graph shows the relative contacts relative to a normal social contacts for the whole population (due to social distancing, but also, additional contact reduction by those exhibiting symptoms, home confinement of suspect cases, and quarantine of detected cases). The graphs show that whereas all countries have reduced their contacts, some more extensively, earlier, or faster than others, affecting the gain of the balancing feedback loop B3 in Figure 5 in different ways. For example, while South Korea was earliest to respond (low threshold), France and Italy showed the largest reduction. Sweden forms a notable exception here by having reduced contacts across all the population by just under 70%. The low early, but subsequent rising value of South Korea and to lesser degree Germany illustrate their (successful) social contact reduction strategy, being aimed at isolating positive cases. Once active case counts go down, symptomatic and suspect cases reduce as well, leading, combined with some general social distancing relaxation, to an increased average social contact rate. Parameter estimates discussed above are consistent with this explanation. Infectious contacts together with transmission delays (incubation time – the time before symptoms begin to appear - and duration of infectivity - of the symptomatic population) determine how many people an infectious person infects during infectivity, affecting the likelihood and extent of the epidemic outbreak. The effective reproductive number R captures how changes in transmission parameters such as social contacts (as well as changes in the remaining susceptible population, negligible here) affect the growth rate of the active cases and thus of the outbreak over time. Because of the relatively high remaining social contact rate their current effective reproductive number is about 3 times that of Italy or France and close to one. Because of this, new cases, and new case detections, remain high (Figure 7, centre right, showing new case detections). While most countries within the selection have for the most been able to deplete the stock of undiscovered cases, for Sweden this is not the case (Figure 7, bottom right, showing the stock of simulated undiscovered cases). Hence, cumulative cases keep growing (Figure 7 left, bottom). While there is uncertainty about the true ratio of actual versus reported cases, the results suggests that the share of the cumulative infections remain magnitudes below values needed for herd immunity (estimated to be about 70% (Ferguson et al. 2020); in a simulation of this the model without interventions and testing, herd immunity builds at cumulative cases of population of 83% people). The case of Sweden suggest that controlled mitigation, if there are no near future opportunities to immunize the population, or a multitude of subsequent outbreak waves, would be very unlikely.

**Figure 7.**
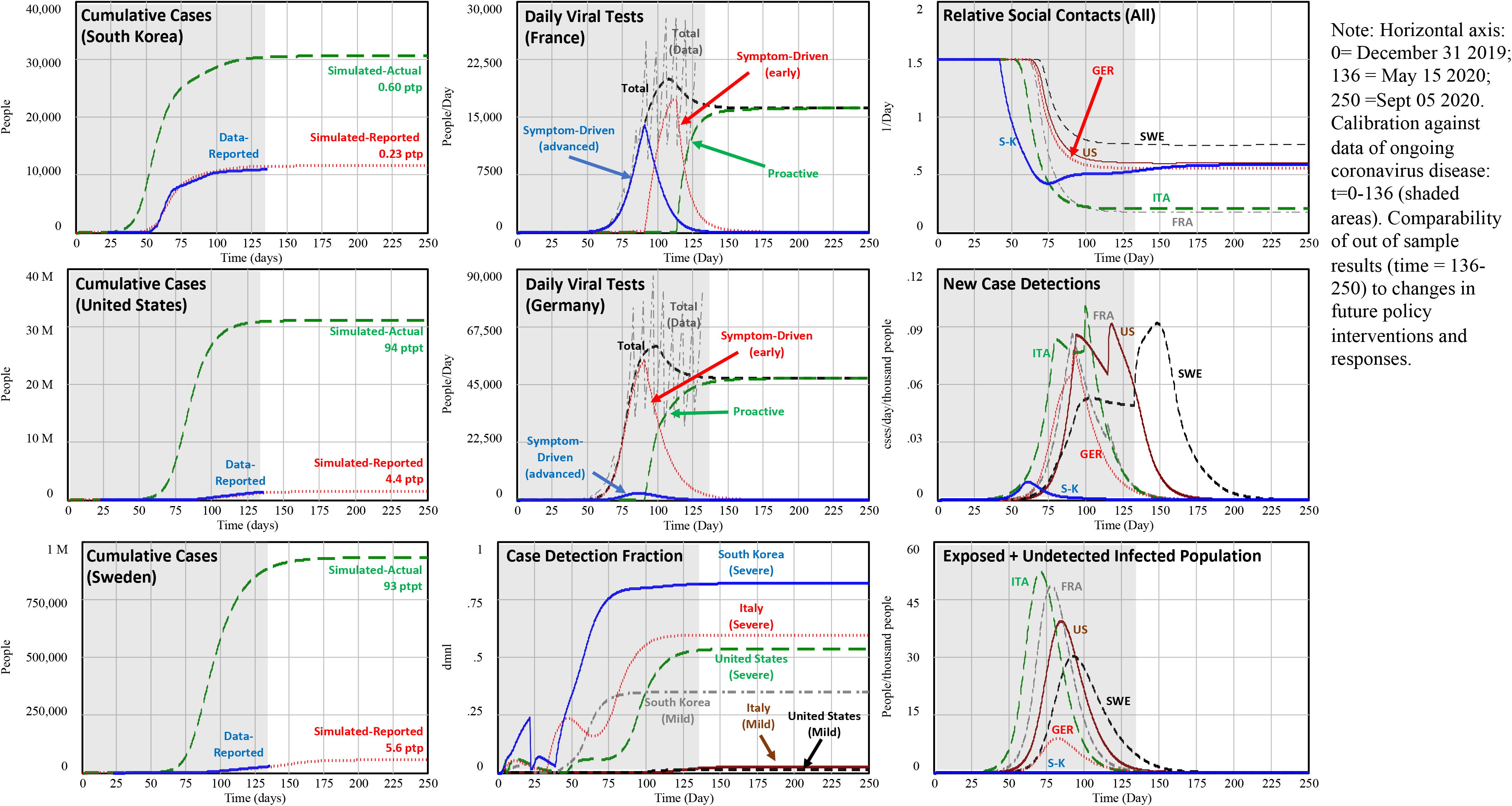
Baseline simulation: Calibrated simulation plus continuation with counterfactual out of sample scenario after May 15 2020 assuming that social contact reductions are maintained until final time.

### Experiment 2 – Sensitivity of baseline results to behavioral responses

We next examine the effect of hypothetical changes in policy and citizen responses to the outbreak. We begin by using the Baseline for the United States as a reference from which we alter three distinct parameters and perform sensitivity analysis of simulated actual deaths (and reported and actual cumulative cases) to changes in: i) the reference outbreak level for policy response (*o_ref,US_*, indicated as RI); ii) maximum contact reduction fraction through general social distancing (*sd_max,US_*, MSD); iii) the threshold for testing capacity the cumulative hospitalization for testing growth rate (though *rCH_US_*^*^, RT); and, iv) their joint effect (All). Figure 8 (left) shows a bar graph of the simulated actual deaths (t=250, 5 September 2020) resulting (“High” and “Low”) the values used for the parameter settings. They values roughly correspond to high (low) policy/citizen responsiveness to the outbreak as observed across different countries in the sample. (Table A.3 shows the parameter values, to be compared with the Baseline values in Table 3). One can see that a more responsive government to the outbreak, citizen’s social distancing, and earlier testing ramp up all have the effect to reduce cumulative deaths. Hence, the results suggest that each of the policy measures taken – earlier and more extensive can drastically reduce the outbreak. On the other hand, any reduced responsiveness greatly exacerbates the outbreak. Further, we see a strong interaction effect among socio-behavioral responses (See “All” vs individual changes).

**Table 3.** Estimated values of model parameters varying across countries. Values are rounded for visual purposes.

| Country | S-K | GER | ITA | FRA | SWE | US |
| --- | --- | --- | --- | --- | --- | --- |
| **Virus Transmission and Clinical** |  |  |  |  |  |  |
| Relative Fraction Actual Infected Cases Severe<sup>3</sup> $rfs_d$ [dmnl] | 1.5* | 0.7* | 0.73 | 0.97 | 0.70 | 1 (ref) |
| Relative Infection Fatality Rate (Severe)<sup>4</sup> $rifrs_d$ [dmnl] | 0.99 | 1.22 | 1.5* | 1.394 | 1.5* | 1 (ref) |
| Time at total cumulative transmissions exceed 100 $\tau_{Ed}$[days] | 21.9 | 21.5 | 1.6 | 9.1 | 34.5 | 5.1 |
| **Social Contacts** |  |  |  |  |  |  |
| Reference Outbreak Level, $o_{ref,d}$ [dmnl] | 0.023 | 1.10 | 2.44 | 0.81 | 0.18 | 2.37 |
| Maximum Contact Reduction Fraction (Social distancing) $sd_{max,d}$ [dmnl] | 0.595 | 0.628 | 0.862 | 0.881 | 0.489 | 0.596 |
| Contact Reduction Exponent (General; Sympt - Relative) $\beta_{sd,d}; rcrs$ [dmnl] with $\beta_{crs,d} = rcrs \cdot \beta_{sd,d}$ | 1.53;0.02 | 0.19;0.16 | 0.30;0* | 0.21;0* | 0.39;0.08 | 0.23;0.15 |
| Relative Efforts Needed for Full Home Confinement Ability $efahc_d$[dmnl] | 0* | 100* | 100* | 100* | 100* | 100* |
| **Testing** |  |  |  |  |  |  |
| Relative Testing Capacity Growth Rate $gr_d$ [dmnl] | 3* | 2.05 | 1.12 | 1.52 | 0.76 | 1.51 |
| Relative Cumulative Hospitalization for Testing Capacity Start $rCH_d^*$[dmnl] | 0.264 | 0.326 | 1.31 | 8.42 | 0.98 | 3.92 |
| Desired Relative Testing Capacity $rtc_d^*$ [dmnl/day] | 0.20e-3 | 0.73e-3 | 1.08e-3 | 0.32e-3 | 0.62e-3 | 0.65e-3 |
| Relative Targeted Testing Needed for Full Targeted Testing Ability $efata_d$[dmnl] | 0.03 | 100 | 0.32 | 0.86 | 0.84 | 0.43 |

<sup>3</sup> The fraction of severe cases is expected to vary, to some degree, across countries. Therefore, with $fr_d = rfs_d * fs$ , with $fs$ listed in Table 2 and $rfs_d$ estimated using a limited range (0.7-1.5%) while setting $d=US$ as reference ( $rfs_{us} = 1$ ).
<sup>4</sup> Infection mortality rates are expected to vary, to some degree, across countries, even controlling for severity of cases. Therefore, with $ifrs_d = rifrs_d * irfs$ , with $irfs$ listed in Table 2 and $rifrs_d$ estimated using a limited range (0.7-1.5%) while setting $d=US$ as reference ( $rifrs_{us} = 1$ ).

**Figure 8.**
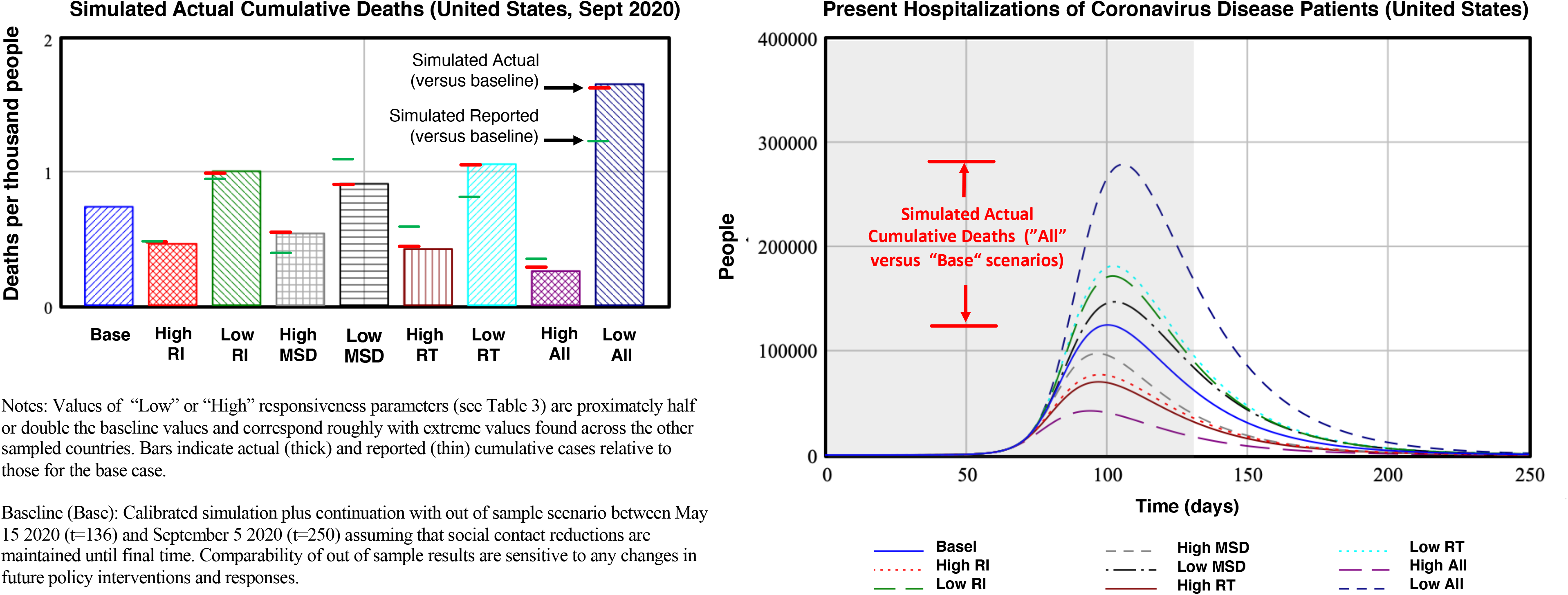
Left Panel: Sensitivity of Cumulative Actual Deaths directly attributable to COVID-19 (*AD_US_*) to changes in policy and citizen responses compared to the baseline (Base): Responsiveness of interventions (RI) to the outbreak, maximum social distancing (MSD), responsiveness of testing capacity (RT), and their joint effect (All). Right Panel: Sensitivity of present hospitalizations of Coronavirus Disease patients to same parameter changes.

Also indicated in the bar graph are simulated actual (thick lines) and reported cases (thin lines) compared to the Baseline results. While actual cases correlate highly with the actual deaths, reported cases are particularly less responsive in the test (RT) scenario because High (Low) responsiveness implies that higher (lower) testing capacity makes up for the increased (reduced) actual cases of which more (fewer) are captured. Further, increased actual cases creates precisely those problems that make it hard to keep up with testing. This observation is important because reported cases are main drivers for decisions and citizen responses. This itself contributes to the strong effects we observe in the actual cases. While it is problematic to use responsiveness to reported deaths as indicator (with lags between infection and death being 3-4 weeks), the sensitivity analysis shows the risk of underestimating the effects of too little action, when driven by reported cases (especially when relative reported cases are low).

The sensitivity to timing and extensiveness of interventions has not only implications for long-term indicators such cumulative deaths, but also affects transitory variable, with implication on the health system. Hospitalizations up build with long lags between transmission, onset, and appearance of advanced-stage symptoms. Hospitalizations decline after recovery or deaths which itself can last several weeks. Together this implies strong amplification of peak hospitalizations (Figure 8, top right, with response to same policy parameter changes). The scale is comparable with that for cumulative cases (red bars in the figure). However, while the day-to-day changes in cumulative deaths or cumulative cases are relatively moderate (not shown), for a transitory state like hospitalization a similar amplification can build up within 30 days, with dramatic consequences for manageability of the health system.

Figure 9 further highlights the strong interaction effect between behavioral responses to the outbreak. Figure 9 (left) shows the joint sensitivity of actual cumulative deaths (*AD_US_*, per thousand people) to changes in reference outbreak level for policy response (RI) and the threshold for testing capacity the cumulative hospitalization for testing growth rate (RT). Higher responsiveness to interventions (RI) is towards the left and to testing (RT) is towards the bottom (Table A.2 indicates the parameter values.). In the graph a number of reference points are indicated with values for *AD_US_* (darker colors indicate higher deaths), including those corresponding with the Baseline (*AD_US_ =* 0.80) and the Low/High univariate changes in Figure 8. Also indicated are simulated initiation times of interventions and of testing buildup for the Baseline case 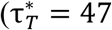 and 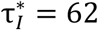) as a result of the responsiveness thresholds, with differential values, compared to the Baseline, for the other reference points. The results highlight the strong sensitivity to the timing of the buildup of testing capacity (top to bottom), corroborating the conclusions inferred from the different baseline results Figure 7 (center graphs). Additionally, however, while a moderate higher/lower response has the effect of reducing (lighter colors)/increasing (darker colors) cumulative deaths, their joint effect amplifies these impacts. Thus, for example, policies that stimulate social contact reduction efforts are greatly enhanced when policymakers and citizen have a more accurate perception of the extent of the outbreak. Compare for example the two red dots (Low RI, *AD_US_* = 1.06 and Low RT, *AD_US_*=1.10) with the white dot indicating the joint effect (Low RI & TI, *AD_US_* = 1.44). Note that the delay in the intervention response compared to the Baseline is larger for the joint effect of RI and RT than for RI alone 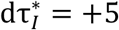 versus 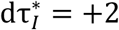. The other way around (towards high RI and high RT) works in the same way, with strong synergistic effects. While at this point care must be taken in taking too much confidence in specific parameters and trajectory outcomes, quantitatively the results suggest that higher (lower) responsiveness in either testing capacity buildup or of social distancing resulting in accelerated (delayed) action by a week could reduce (increase) cumulative deaths by about 50% (about 0.5 deaths per thousand people). Combining responsiveness in intervention responsiveness could further reduce deaths.

Figure 9 (right) further highlights the importance of addressing all policy dimensions for to be able to suppress the outbreak. The Figure shows, now for South Korea, again the effect of the reference outbreak level for policy response (RI) on actual cumulative deaths, but now jointly with the effectiveness of one of targeted approaches - the Ability to Identify Associated Cases for Home Confinement (SUS). Higher responsiveness of interventions (RI) are again towards the left and towards the top for SUS; the color coding is rescaled to match much lower deaths for South Korea. The Baseline result is indicated in the top right. The sensitivity analysis for South Korea suggest that, while weaker responses in either RI or SUS have large relative effects on deaths, they do not alter the scale of the epidemic impact. This is so because South Korea had a number of policies in place that enabled it to respond effectively beyond rapid response and home confinement of suspect cases this includes extensive testing and contact tracing home. The slack in testing capacity frees up resources for proactive testing and helps build up a stock of potentially identifiable existing and future cases. Thus by taking these multiple actions timely and aggressively they created a critical redundancy in response to the outbreak. The analysis suggests that, for the same reason just improving testing once you fall behind - such as the case for the United States, is not sufficient.

**Figure 9.**
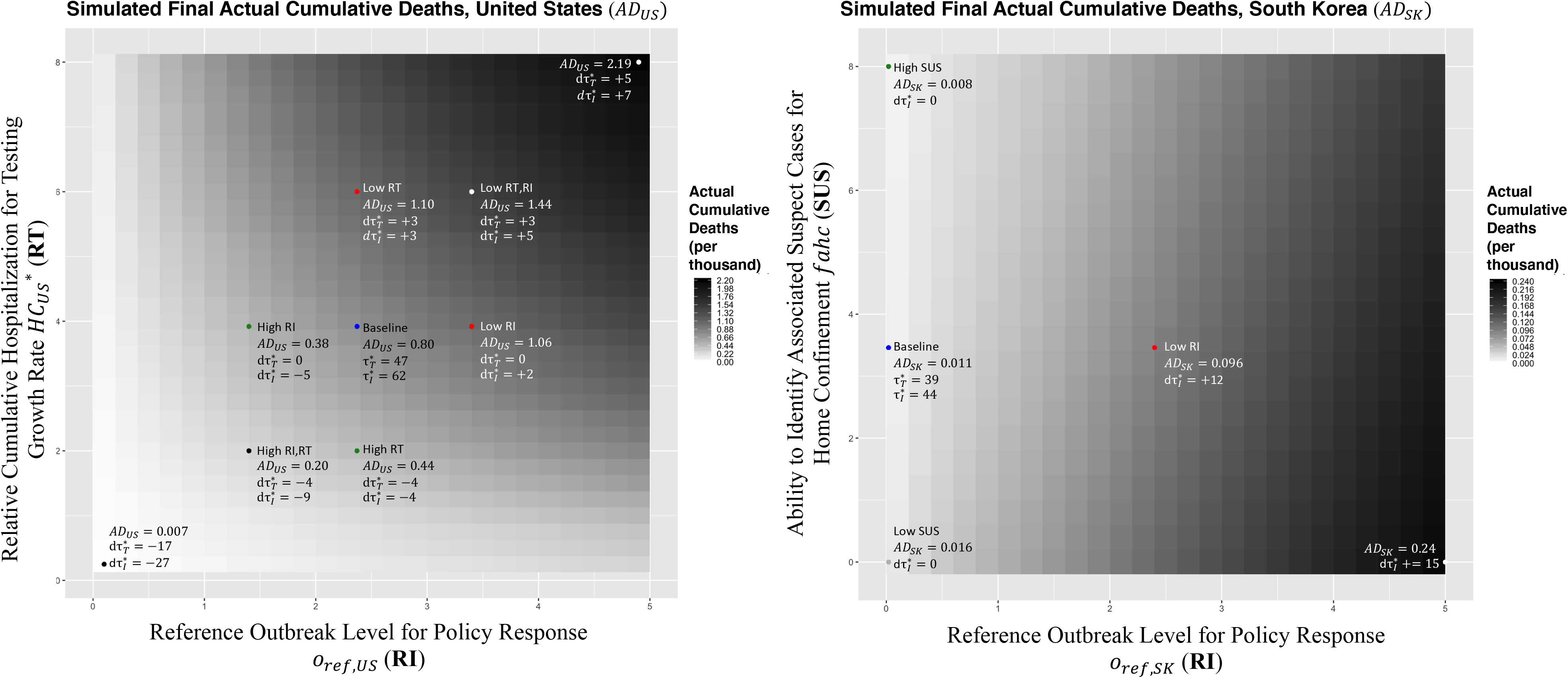
Sensitivity of cumulative deaths to policy interventions and citizen responses. Left: Interaction between responsiveness of interventions (RI) and testing capacity buildup (RT), for the United States. Right: Interaction between responsiveness of interventions (RI) and ability in identifying associated suspect cases for home confinement (SUS), for South

Together the results highlight together the extraordinary measures in many of the early outbreak countries were critical to control the outbreak. In particular the combination of testing and finding ways to reduce general social contacts are critical. More targeted approaches can work as long as complementary resources (identification ability, testing, surveillance) and slack in testing capacity are available.

### Experiment 3 – Managing Deconfinement

The following experiment focuses on the challenge of managing deconfinement. Illustrating one aspect, we define deconfinement here as the reduction (to some degree) of social distancing for the general population. Because confinement involves high social and economic costs, there is large pressure to reduce this at some point after (or even before) active cases begin to reduce. But how much can a country deconfine? What system needs to be in place in a country to be prepared for deconfinement? Specifically, how do targeted interventions and testing help the deconfinement? To illustrate some of the key tensions we perform an analysis of a stylized context. The stylized context, while perhaps harder relatable to a specific real-world case, allows for better comparability across results and, through that, easier develop an internally consistent and reliable explanation of the results. In developing the setup we stay as much as possible to the country cases we have examined. Consider a region of 16M people (the approximate size of a metropole like New York or Paris, of the hard-hit region of Northern Italy, or of a country like the Netherlands) with as starting point characteristics similar to the average of the countries we analyzed before in the Baseline (see again Table A.3 for all the parameter settings). We then introduce a COVID-19 outbreak with 100 undetected infections. Next, at deconfinement time *τ_dc_*=10, counting from the first time reported new cases begin to decline, policymakers begin to reduce a fraction *fdc* ∊ [0,1] of the social distancing that has endogenously emerged. A value of *fdc =* 0 indicates no intervention (similar to our counterfactual Baseline scenario) while *fdc =* 1 means full deconfinement. Deconfinement is ramped up to the level indicated by *fdc* during a period *τ_dd_* = 60 days after which the deconfined state remains at that level. Deconfinement is restricted to contact reduction policies related to undetected and non-suspect cases only – so, for example, quarantining of detected cases and home confinement of suspect cases (see Figure 2, 5) remain in place. We perform the experiments, varying a number of parameters with particular focus on the role of targeted approaches during deconfinement.

Figure 10 shows, as before, a number of graphs of simulated cumulative actual deaths *AD*, at time=500, here as a function of the ability to identify associated suspect cases for home confinement *fahc* (SUS, horizontal axes) which we also used in the previous experiment (Figure 9), and the deconfinement fraction (FDC, vertical axes). We further vary maximum social distancing prior to deconfinement (MSD: left panel (Low) versus right panel (High)), as well as contract tracing ability (CT, within each panel: left ((Low) versus tight (High)), and the relative initial experience with targeted interventions and testing (ETA: top (Low) versus bottom (High)). Table A.3 lists again all the parameters adjusted for the experiment. Each graph shows two reference points with the at zero deconfinement efforts (FDC=0) indicating *AD* for respectively zero and high ability to identify suspect cases for home-confinement. The graphs indicate parameters regions with actual deaths exceeding 2 per thousand (*AD*>2), a number greater than the largest number of deaths absent deconfinement (bottom left of each graph, FDC=0 and SUS=0).

The graphs show, first, the very strong threshold as a function of the deconfinement fraction. Simply put, above this threshold, the reproduction number becomes larger than one (R>1) and the outbreak keeps growing. Thus, avoiding deconfinement risk requires well below this threshold. The different figures show what conditions and measures and combination help doing this, or, which alter this threshold. Across the graphs one can see that a strong ability to identify and home-confine suspect cases can help increase the level of deconfinement at which R stays below 1. The contrasts between the bottom graphs (high initial experience with targeted approaches) versus the top graphs (low initial experience) show that having abilities to identify and isolate suspect cases needs to be in place before deconfinement starts. Once deconfinement starts, active cases decline at lower rate than otherwise, or may grow again, or, and with that, resources to perform targeted approaches as well as general testing may become overloaded. Therefore, countries such as South Korea and Germany are expected to be able handle deconfine the general population further than most other countries. Second, comparing the left versus right graph within each panel suggests some positive interaction between contract tracing ability (CT) and home-isolation of suspect cases (SUS). By itself contract tracing may not have a large impact on deconfinement outcomes because contract tracing does not directly alter the reproductive number much. However, contact tracing does enable a more effective isolation of a number of potential positive cases and in this way strengthens the effect of suspect case identification. To illustrate this, the benefits of CT are larger for larger SUS than for smaller SUS.

The deconfinement analysis so far suggests the vital importance of capacity for targeted approaches (general testing capacity, effective contact tracing and testing, extensive and effective confinement policies for suspect cases). Next we ask, how is the extent of deconfinement that is feasible affected social distancing policies prior to deconfinement? The right panel (High MSD) versus the left panel (Low MSD) provides a view on this problem. To directly compare the extent of deconfinement, one has adjust the deconfinement fraction for one of them because the same deconfinement fraction yields different absolute MSD post-confinement. The horizontal reference lines shows the same absolute deconfinement of respectively 70% and 80% compared to 100% social interactions of the general population for the low and high MSD case. Once can see that well developed targeted approaches (compare the bottom left panel versus bottom right panels, at high SUS) the threshold for a renewed outbreak is lower for countries with lower initial MSD. This is presumably the case because the larger number of active cases upon deconfinement make it harder to keep down the suspected case pool. Thus, low MSD countries face larger deconfinement risks than high MSD countries. Thus, a country like Sweden will face larger deconfinement risks than a country like France or Italy, assuming identical deconfinement time and abilities for targeted intervention and testing.

Finally, summarizing the results more quantitatively, absent strong capabilities for targeted approaches that allow in-the field case detection and suspect case isolation, restoring social contact rates to above about 60-70% of pre-pandemic levels leads likely to renewed outbreaks (see the top left areas of all graphs). Targeted approaches may allow deconfinement to increase by 10-30% due to early new case detection and isolation.

**Figure 10.**
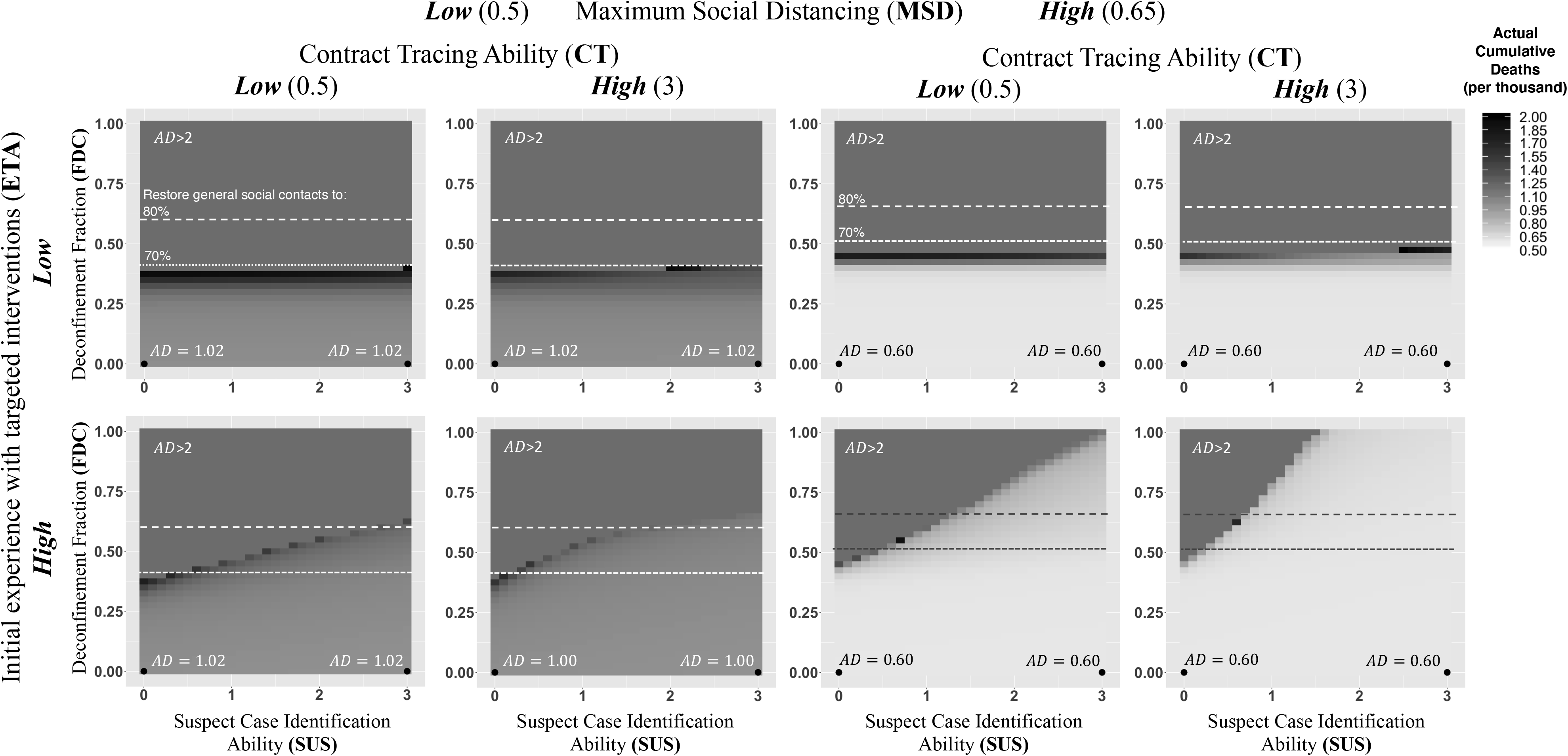
The effect of Deconfinement Fraction (**FDC**) and Relative Ability in Suspect Case Identification (**SUS**) on Cumulative Actual Deaths (*AD*) at time=500. Varying also: Maximum Social Distancing (**MSD**; Left vs Right Panel), Contract Tracing Ability (**CT**; Left vs Right Colum of Left and Right Panel), and Initial experience with targeted interventions (**ETA;** Top vs Bottom Rows)

### Experiment 4 – Managing resurgence

Next, we highlight the challenge of managing resurgence. Because in the short run - without available vaccines - building up herd immunity is not likely a feasible strategy (Ferguson et al. 2020), it is highly likely that additional waves of outbreaks will occur in the near future. Different from a first wave, during resurgence, reasonable testing capacity will likely be available. Hence, in this phase proactive testing approaches (community oriented, and/or contact tracing) will be a feasible as a policy even when not so during the first wave. Within the same setting as for experiment 3, to allow resurgence, we now let policy makers and citizens respond to the reported active (not cumulative) cases. Hence, once citizens and policymakers have begun to relax their social contacts, new cases gradually grow again. Different from the deconfinement scenarios, we now allow society to return to contact reduction strategies in response to a resurgence. Depending on response sensitivities and behavioral and virus transmission delays a cyclical pattern of active cases results. Figure 11 shows simulations using synthetic data of a representative case (See Table A.3 for all parameters differing from the Baseline.) The base case is the blue (continuous) line, that does not include any targeted approach. The simulated actual active cases (top left) show the cyclical pattern. At some point the virus outbreak appears to be receding, as actual (and reported) cases go down considerably. As social distancing rebuilds (right) the virus can transmit again easier among the population allowing a second wave to commence, and so forth. Cumulative deaths (bottom left) keep rising considerably with each wave. While, naturally, susceptible cases decline (inset), the decline is not sufficient to build strongly reduce R, let alone build herd immunity. The cyclical patterns of social contacts and hospitalizations highlight the high societal costs associated with the base case.

**Figure 11.**
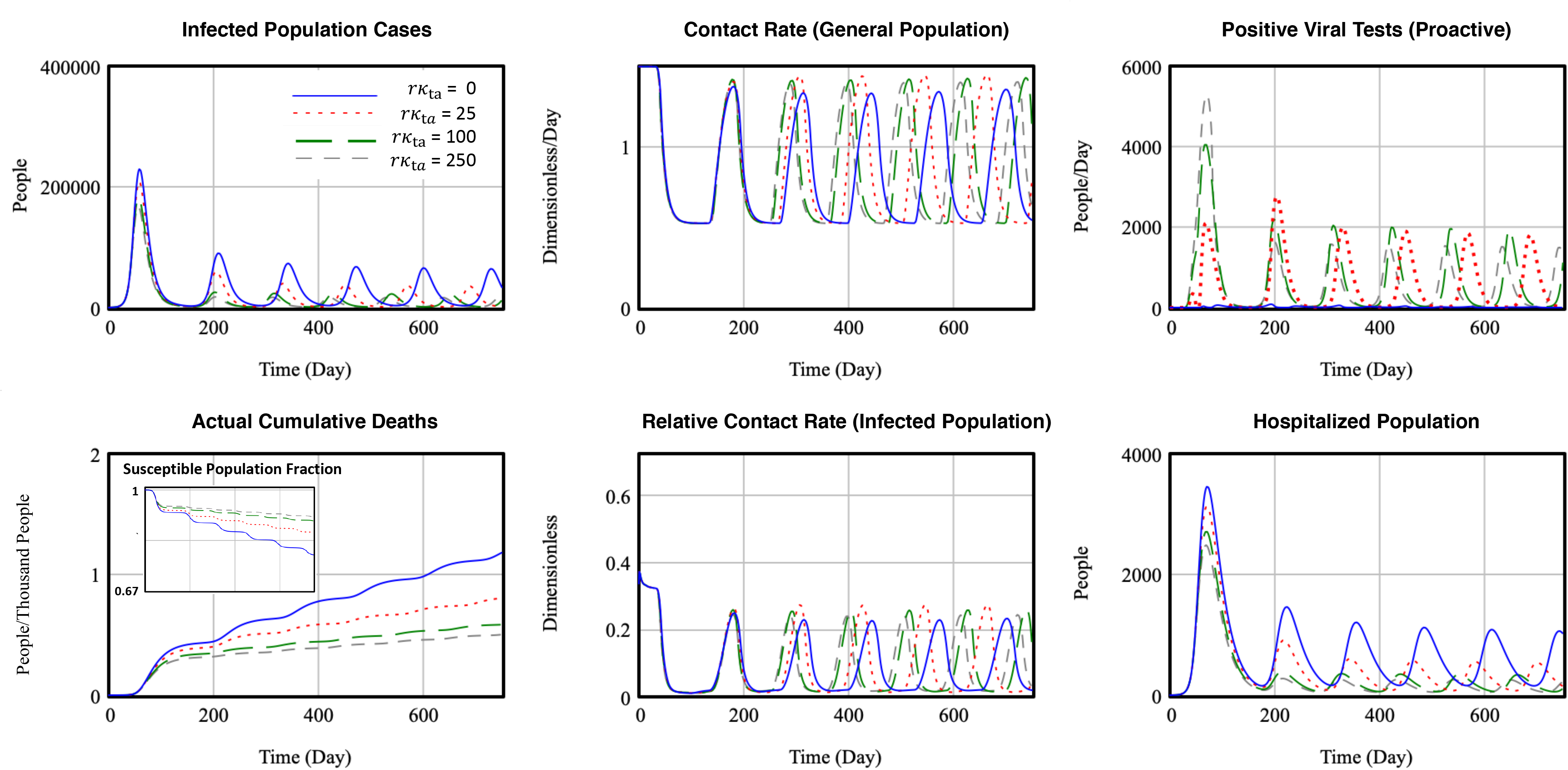
Resurgent outbreaks (response to reported ***active*** cases), varying targeted testing effectiveness (measured by the Relative Targeting Effectiveness Parameter & *rκ*_t_*_a_*)

In this case, proactive testing may be expected to play a critical part of effective responses to a breakout because the society will also respond by reducing social contacts. Having an ability to identify positive cases could help in this responsiveness. To understand the impact of targeted approaches, we now vary targeted testing effectiveness, measured by the clustering parameter *rκ*_ta_. The strongest effectiveness corresponding with *rκ*_ta_=50. First notice that all scenarios respond similarly to the first wave, irrespective of proactive testing effectiveness. This is so because, as before, testing capacity is initially constrained. Therefore, proactive testing can do little to alter the path of the first wave during which testing capacity is being built up (and lags). We note further that this firs wave is similar to what it would have been in case of a population responding to cumulative cases.

While the oscillation resist suppression in the base case, proactive testing effectiveness (higher r*κ*_ta_) is very effective in dampening the oscillation of new cases. The rapid detection of new cases takes an important share of the newly introduced symptomatic population out of contact (bottom right, symptomatic population remains at a relatively low level. The policy is very effective in not only reducing oscillations and overall emergence of new cases, but also, in this case, suppresses oscillations of social distancing and, for high contact tracing, stabilize this at around 60%. Scenarios with lower social distancing sensitivity (not shown here) can more rapidly stabilize social contacts, at higher levels of about 70% of normal contacts. While more work is needed to analyse resurgence dynamics, this analysis suggests that reduction in responsiveness to internally produced resurgence, under circumstances, be more important as this stability helps the effectiveness of targeted approaches that benefit from stable use of testing and other resources.

### Experiment 5 –Dynamics of vulnerable population segments

Symptom severity, hospitalization, and infection fatality rates differ considerably across age groups, with the older population being disproportionally vulnerable to the impact of the virus (Russel et al., 2020; Russell et al., 2020). In a final analysis we use the model segmentation to help better understand how these differences affect the different population groups as well as the overall outbreak dynamics. To illustrate the value of analyzing this in more depth, we focus in particular on explaining the role of different social interactions across these segments in different countries in explaining the outbreak patterns. We perform an analysis in the same stylized way as the previous experiment. Doing this helps focus on the key dynamics at work. We again use a stylized region of 32M (2*16M) people, but now we differentiate the population into two age cohorts, differentiating those that are more and less vulnerable (consider “older” and “younger” populations, though note that the distinction can also proxy other stratifying variables such as income or race.). We control the difference between the segments by varying their relative severe cases (*rfs_d_*), holding the average infection fatality rate constant across the segments. Testing capacity grows as before, with an initiation threshold of 100 hospitalized cases. At time 0 we introduce again a COVID-19 outbreak with 200 undetected infections, but only within the less vulnerable population segment. (Table A.3 shows parameter details.)

Figure 11 shows the results. The graphs show on the horizontal axis the relative cases severe (*rfs*_2_) across the segments (relative to within segment contacts). A value of one indicates that the fraction of severe cases (and therefore the same case fatality rates), is identical in the two segments, while moving to the left signifies a relative severity of cases (and with that case fatality) that increases for the vulnerable population. The left graph shows the actual cases (left vertical axis, as share of the population) and deaths (right vertical axis, percentage of total population in the stylized simulation). We also vary relative contact rates between the segments (*fc_d′d_*). Continuous (dashed) lines have high (low) intersegment contacts. (When varying contact rates between segments we control for total contact rates within the population.) The right graph shows the cumulative death fraction, in three different ways: total versus reported, total versus actual cases, and as the share of vulnerable population to the total. (For reference, one can see that this share reaches to 50% at high intersegment contacts, when *fs*_1_ *= fs*_2_.) The graph shows a number of interesting insights about how cases and deaths develop as we vary these two parameters. In particular, when case fatality is very uneven, actual cases increase (left graph). This is so because low vulnerability implies (mild on average milder symptoms and, because of that, lower detection. With low reporting there is little policy response and infections can easily spread among the less vulnerable populations. A relevant, but not unrealistic starting condition in this analysis is that the outbreak initiated among the young population. For example, in regions like New York City it appears that socially active younger population (with mostly mild symptoms) one can envision that the virus has spread rapidly but fairly undetected for a while. The results further show that the worst case in terms of absolute fatalities, high variation in case fatality, and relatively high contacts between the population segments, disproportionally affects the vulnerable population (dashed line top right.) This is so because after the virus spread has spread among those who are less vulnerable, it can easily spread to the vulnerable population segment, at which point it is uncontrollable. It is the latter that may have be playing part in Italy, with relative contacts across generations generally being larger than in many other countries.

While only a synthetic analysis, this analysis on stratified vulnerability may partially help explain the large variation in reported death fractions (case fatality fraction, CFR) across counties. Or, why the reported CFR in some countries seem low for a while, only to go up later, counter the expectation as recoveries and death accumulate. These insights also may have policy recommendations may vary depending on the demographic makeup. For example, when relaxing general confinement policies, or when managing resurgence, should populations that are deemed more vulnerable remain (longer) isolated at home? Based on the insights here, that may be a feasible direction, though provided that mild cases can be surveillance and isolated.

**Figure 12.**
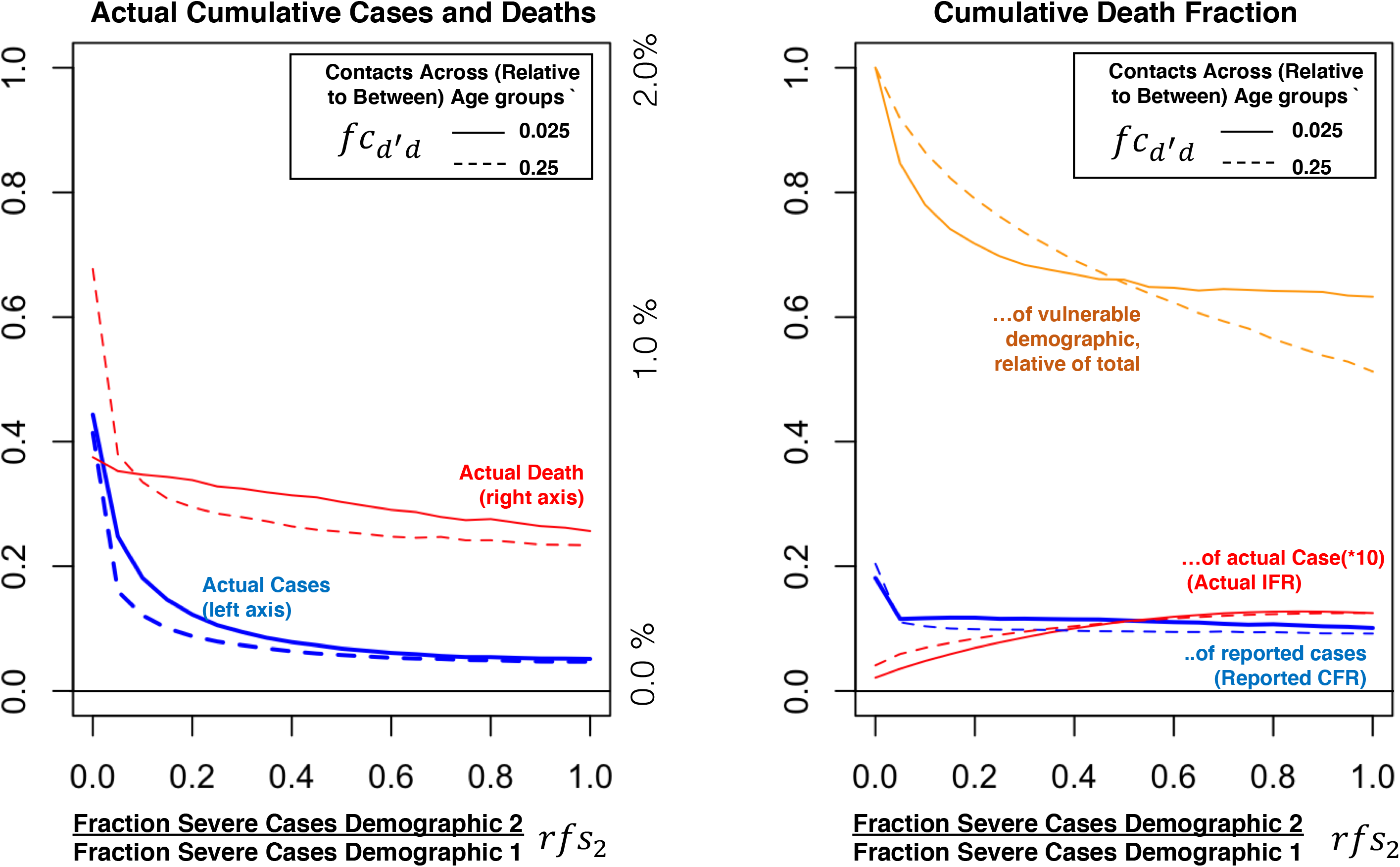
Simulation of outbreak impact as a function of relative severe cases across two (hypothetical) demographic population segments (demographic 1=“vulnerable”; demographic 2=“less vulnerable), also varying relative contacts between segments (populations of both demographics are 50%)

## Discussion

This paper developed a behavioral infectious disease model capturing how virus transmission dynamics and policy and citizen responses interact to shape the course of an epidemic virus outbreak. Applicable to the full epidemic cycle including deconfinement and resurgence, the model allows exploring the impact of individual and joint policy interventions.

To study implications of different interventions and testing strategies the model incorporates some key behavioral aspects and disaggregates critical constructs. Central to the model are not only virus transmission dynamics, following SEIR-based epidemic modeling traditions, but also explicit formulations of policymakers ramping up testing, reporting, and interventions in response to the outbreak, and populations altering their social contacts, with potentially imperfect compliance. Further, the model differentiates between: mild versus severe symptoms of those infected; reactive versus proactive testing; and interventions for general, suspected, and detected populations, through social distancing, home confinement, and quarantining. Finally, the model captures heterogeneity across sociodemographic and geographical population segments, allowing for controlled interactions among them. Impact metrics in the model include reported and actual positive cases and deaths, but also make explicit endogenous social interactions within different segments of the population and hospitalizations. Together these metrics allow consider not only study outbreak dynamics (first wave, deconfinement, and resurgence) but also help consider societal implications - health system overload, recurrent social distancing - resulting from these different scenarios. Thus, the model can be used to evaluate the broader impact of diverse (in particular non-pharmaceutical) public health control measures, to consider interaction with testing and reporting, and citizen response.

Using the model to explore both current questions about managing the COVID-19 outbreak and future questions about managing resurgence yielded important insights relevant to policy and to infectious disease modeling. In terms of policies, I demonstrated the interactive effects of distinct policies and/or of citizen behavior and policies. Efficacious responses to the crisis does not only require willingness to implement general and develop targeted responses but also early testing capacity and ramp up that measure the impact. For example, even under high willingness, limited testing capacity implies delays in implementing such policies. In the case of COVID-19, due to the strength of the transmission feedback and dynamics towards reactive approaches, a one-week delay in testing/intervention buildup can be as costly as 25-50% additional (fewer) cumulative deaths. Second, deconfinement analysis shows that society will have to find ways to effectively reduce physical contacts to at least 40%-20% below pandemic levels as long as no pharmaceutical solutions are available at scale. Confinement policies prior to deconfinement and a country’s preparedness for deconfinement have impact risks of renewed large-scale outbreaks. The presence of effective and extensive targeted testing and intervention approaches (contact tracing and testing, broader suspect case isolation, surveillance, and their compliance) are vital to reduce these risks. Longer interactive dynamics of resurgence and key policy levers highlighted similarly, the importance of targeted testing.

The importance of such targeted approaches raises important questions about how this can be achieved. First, while slowing the rate of deconfinement may not directly reduce R below 1, its pacing may make it easier to monitor and respond to increasing outbreaks and building further capabilities to do so at higher levels of deconfinement. Further, absent guarantee of near-term vaccination or other forms of mass-immunity, societal innovation towards living at 50-70% of physical contacts is critical. Finally, implementing targeted approaches may mean on the one hand reliance on contact tracing apps. Doing this without safeguarding privacy or coordination across countries, this may lead to unintended consequences for society. On the other hand, the need for targeted approaches suggest a strong opportunity to put more responsibility with citizens within communities and support their capability development and awareness for suspect case identification and isolation.

Our final analysis demonstrated the value of strategic disaggregation to generate important insights – such as inequality issues that affect both segments overall outbreak dynamics. Together, the analysis shows the nature of non-linear and multi-feedback system being resistant to change. Our analysis shows what is generally true for complex dynamic systems: Significantly altering the pathway of a focal variable within the system requires a mix of interventions is required to address different positive feedback loops and delays within the system.

While many epidemic policy models exist, this model contributes by combining a range of policy and testing instruments and behavioral aspects critical to infectious diseases with high reproductive numbers, high fatality, but generally low severity of cases. Calibration across a variation of countries with different policies demonstrates its robustness for the case of COVID-19. Further, while dedicated to COVID-19, the model is flexible in that it can be applied to other infectious disease contexts. First, while the analyses demonstrate that fundamental insights can be derived with a relatively aggregate model, additional subsequent empirical analysis on country-or region-level analysis, involving varying epidemic pathways and policies can provide more confidence in the specific parameter values related to both virus transmission and social behavior. The detailed documentation and model accessibility allow replicating and expanding this for similar and other contexts. The operational details and behavioral aspects also allow using it as a policy tool. It is clear that successful interventions require broad support across health exports policymakers, volunteers, citizens, and media - of outbreak control efforts. As part of this, the model is also accessible in the form of a free web-based management flight simulator (Struben, 2020). Doing this enables users to explore the impact of government and citizen responses, and how they could alter the course of a pandemic. Accompanying graphs display the results immediately, including actual and reported people infected, recovered, and deceased, new infections, effective contact rates, and hospitalizations. Users can create different scenarios by altering assumptions about each of these factors, and then create and compare multiple scenarios. Sliders allow users to simulate policy choices and citizen behaviors - for example, how rapidly citizens alter their contacts with others voluntarily (such as staying at home), or adjust government policies on social distancing (recommended versus forced closures), quarantine (targeted versus general), or case testing and reporting. Users can also vary a range of assumptions about the disease transmission parameters (infectivity, contact rates, incubation time, duration of infectivity), or alter the regional characteristics (population size, interregional contacts).

The model and analyses suffer from usual limitations as well as from those related to the emergent case of the outbreak. First, the relatively aggregate representation of population segments implies that important dynamics may be missed. For example the emergence of superspreader events are expected to important in the spread of many infectious diseases (Lau et al. 2016). While the model allows analysis of targeted approaches the model is currently not well equipped to examine such heterogeneity. Likewise, given the relative aggregation in the current analysis, the model likely underestimates the value of targeted approaches. The model in its present form also leaves out important structural elements, such as endogenous infections and fatality within the health-providing system (Fiddaman 2020).

The analyses and limitations listed here suggest at least three clear directions for further work. First, in terms of problem orientation, given that in the current pandemic countries increasingly begin to reach the peak of the first outbreak wave, future analysis should focus on managing the transition towards deconfinement and resurgence waves. The experiments show that the model is well-suited for such analysis. The full model also has a tentative sub structure that allows the implementation and roll-out of (potentially imperfect or slow) vaccination as well consider the role of immunity loss rate. While switched off for the purpose of this paper, over the next year(s) understanding these dynamics in combination with resurgence dynamics will be important. Finally, in cases such as these, with need for on the ground learning within a turbulent and dynamic environment and with limited and emerging data, it is critical to have a tool that allows investigating and provides sufficient clarity so that it can forms the basis for policy discussions that are grounded in science and in formal representations whose behaviour they produce can be explained – and then challenged and/or build upon - in internal consistent ways.

## Data Availability

all data is publicly available (sources have been referenced in the document.)

## Acknowledgements

I am thankful Tijn Struben-Huising for support work on the visualizations, to helpful comments and guidance from the journal editor and reviewers.

## Footnotes

1 In addition the model can be used to explore how the dynamics change depending assumptions related to infectious diseases, such as SARS, HIV, H1N1, and H5N1, Influenza, and Ebola.

2 Most of those countries/regions that responded well have had earlier experience with the SARS epidemic (2002–03).

3 Reported cases and testing data were retrieved from Ourworldindata.org (Roser et al. 2020). The metric for social contacts is a composite of data on intensity of walking, driving, and public transport use, relative to the reference of January 18 2020 (retrieved from the Apple Maps Mobility Trends Reports, Apple 2020), each contributing 1/3 to 50%, and of data on school closings (retrieved from Unesco, 2020), one a relative scale from 1 (all schools open) to 0 (all schools closed), contributing to the remaining 50%.

4 The Technical Appendix is available in the online Appendix folder: https://www.dropbox.com/sh/x35h4t8mkkybe0v/AAAkBGr0iWlo6lN3tVpmIb-Va?dl=0. The Appendix folder also includes the full model (in Vensim software), the data used for the calibration analysis performed in the paper (as well as for the selection of other regions and countries), and all files necessary for replicating the analysis. I explain the data, calibration process, and estimated parameters here briefly.

5 This is a simplified representation of the virus transmission rate. In the model the stocks of exposed and symptomatic populations are each disaggregated into different stocks with different contact rates. For example, part of either population segment may be quarantined or home-confined(discussed below).

6 Different formulations have to be considered here. The linear formulation is a special case of a more robust non-linear weighed formulation of the reported cases and deaths. However, this would introduce additional parameters. Such a richer formulation is most relevant when the ratio of deaths and cases vary over time, or when comparing across different virus/diseases with varying infection fatality rates across them. For our purpose the simpler formulation satisfies. The model uses the general classic CES formulation (McFadden 1978) but then sets this to the linear form. Regarding inputs, an alternative is to use recent deaths or active cases to be able to capture easing of social contact reductions as the perceived severity of the outbreak declines. However, such formulation would likely have to be more complex. Moreover testing the effect of deconfinement interventions in controlled ways the above formulation using cumulative reported values is preferred. Further, in formulation In experiment 4 below examining resurgence uses an alternative formulation of active cases (Table A.3).

7 In the full model the prioritization is not necessarily purely hierarchical. A parameter “sensitivity for testing prioritization”, *β_t_* indicates the propensity to allocate all necessary testing capacity according to priority versus along relative desired testing rates. *β_t_*=0 implies tests are performed according to capacity needs (and hence, implies a tendency away from reactive testing). Higher values implies stricter allocation according to priority, with in the extreme tests capacity being allocated strictly according to priority (implying a tendency towards reactive testing). The simulations below run with fully hierarchical allocation of testing capacity.

8 The Appendix (see Model section) folder includes all files and data necessary for replicating the experiments including the calibration, using the provided Vensim model. Appendix II describes the estimation procedure and data in more detail. Appendix IV includes the instructions for replicating the experiments.

9 A note of caution: Any simulation, but in particular out of sample simulations during a developing case (as this one) should be interpreted with great caution and scepticism, for multiple reasons. First, confidence bounds are not indicated. This is partially because of visual clarity but also because there are many structural assumptions (such as about the relative weight of data types) that make this less meaningful. There is still much uncertainty about many of the transmission parameters. Further, estimation of behavioural parameters is done at an aggregate level and therefore simplifies the cross region diffusion. This is particularly relevant for the United States. Finally, forward looking outcomes do not incorporate future policy and citizen actions that may different from those simulated here. Yet, the uncertainty is fundamental to the problem itself. The main purpose here is to develop and enhancing a grounded understanding of the problem, to build confidence in and allow challenge what are brought forward as plausible explanations, and support in the best way those policy decisions that need to take place under this great uncertainty.

